# Widening Ethnic Inequalities in Stroke Incidence: A 30-Year Population-Based Analysis of the South London Stroke Register

**DOI:** 10.64898/2026.01.13.26343995

**Authors:** Camila Pantoja-Ruiz, Amal R. Khanolkar, Ismail Ismail, Aliya Almirova, Evelyn Lim, Marina Soley-Bori, Wasana Kalansooriya, Eva S. Emmett, Abdel Douiri, David Wyatt, Yanzhong Wang, Rachna Chowla, Ajay Bhalla, Anthony G. Rudd, C D A Wolfe, Matthew DL O’Connell, Iain J. Marshall

## Abstract

**Aim/background:** Ethnic inequalities in stroke incidence are well-established, but long-term trends and the impact of socioeconomic status (SES) remain understudied. We estimated incidence rates by ethnicity over 30-years, and differences by SES, using the South London Stroke Register (1995–2024).

**Methods:** We conducted a population-based study including all residents of a geographically defined area of South London, calculating standardised incidence rates using census-derived denominators and the 2013 European Standard Population. The area-level Index of Multiple Deprivation was the indicator of SES. Poisson regression estimated incidence rate ratios comparing ethnic-minority vs White groups, adjusting for age, sex, and SES, with ethnicity-SES interaction testing and marginal rates derived by subgroup.

**Results:** Among 7,726 people with first-ever stroke, overall standardised incidence decreased 34% between 1995–1999 and 2010–2014 (198;[187.8–208] to 131;[122.4–138.9] per 100,000), then increased 13% in 2020–2024. Black-African and Black-Caribbean populations had higher baseline incidence persisting across all periods. Ethnic inequalities narrowed initially but subsequently widened, with incidence twice as high in Black-African (IRR=2.31;[2.03–2.62]) and Black-Caribbean (IRR=2.00;[1.73–2.31]) groups compared to White groups in 2020–2024. Patterns were consistent across stroke subtypes, with the largest inequalities in primary intracerebral haemorrhage. Ethnic differences in incidence attenuated on SES adjustment but remained significant. Within ethnic groups, most deprived Black-African and Black-Caribbean populations showed highest incidence.

**Conclusion:** Stroke incidence has risen sharply in the past five years, driven by rising incidence rates in Black-African and Black-Caribbean populations, with reduction in the White population. Achieving equity requires ensuring uptake of cardiovascular risk programmes across all socioeconomic and ethnic groups, with earlier risk management for groups who develop stroke 10-12 years younger than the general population.

## Introduction

Stroke remains a leading cause of death and disability worldwide^1^ with persistent ethnic and socioeconomic inequalities despite advances in prevention and care^2,3^. In high-income countries (HIC), minority ethnic groups experience higher and earlier incidence than White populations^4–9^. In the US, Black adults have incidence rates 1.5-2 times higher than White adults, particularly at younger ages^4^. In the UK, previous studies using the South London Stroke Register (SLSR) documented similar ethnic inequalities, with higher incidence in Black individuals further pronounced in younger adults ^6,7,9,10^. Globally, the GOAL meta-analysis found higher incidence and risk factor prevalence in Black and Asian patients, with higher mortality in low- and middle-income countries (LMICs) despite younger age^11^.

These inequalities are driven by multiple factors^12^: higher prevalence of hypertension^13,14^ and type 2 diabetes^15^ among Black populations, compounded by socioeconomic disadvantage, healthcare-access barriers, environmental exposures including air pollution^16^, limited green space^17^ and psychosocial stressors^18^. Population-level improvements in hypertension diagnosis, control and smoking reductions, have not been equally achieved across ethnic groups, contributing to persistent and widening inequalities^10,19,20^.

Long-term data on stroke incidence by ethnicity are scarce, requiring sustained case ascertainment, consistent follow-up, and adequate minority group representation; all resource-intensive^21^. Whether ethnic inequalities have narrowed over time remains uncertain. Socioeconomic circumstances vary between and within ethnic groups, and the extent to which socioeconomic status (SES) explains ethnic inequalities is unclear. Hospital-based evidence shows that cardiovascular outcomes and other chronic diseases as cancer differ by both ethnicity and SES, with studies as Myocardial Ischaemia National Audit Project^22^ and HELIUS^24^ showing worse outcomes in minority ethnic groups and more deprived populations^22–24^

The SLSR, established in 1995 as a population-based register covering a multi-ethnic inner-city population, provides comprehensive long-term surveillance with detailed case ascertainment, and multiple SES indicators^25^. Previous studies reported ethnic inequalities in stroke incidence, with declines in White but not Black populations over 10–16 years^6,7,9^. However, these analyses were limited by shorter follow-up periods and did not examine the impact of SES^6,7,9^.

With three decades of data now available, this study estimated standardised stroke incidence over three decades, overall and by ethnic group, stroke subtype, and sex. We further examined whether ethnic differences persisted over time and whether SES explained them.

## Methods

### Study Design

The SLSR is a population-based cohort study with prospective recruitment of all people with first-ever stroke from a defined geographic area in South London since 1995^25^. Ethical approval was obtained from the NHS Health Research Authority and ethics committees of Guy’s and St Thomas’ Hospital, King’s College Hospital, King’s College Hospital, Queen’s Square, Wales REC 1Research Ethics Committee, and Westminster Hospital. Written or verbal informed consent was provided by all participants or their legal representatives. This study adheres to the Strengthening the Reporting of Observational Studies in Epidemiology (STROBE) guidelines.

The SLSR uses multiple-source case ascertainment for all first-ever strokes occurring among residents of two London boroughs, Lambeth and Southwark^25^. Since 1995, trained study personnel including physicians, nurses, therapists and non-clinical fieldworkers have conducted daily and weekly surveillance of all local hospitals, including acute stroke units, emergency departments, general medical wards, neuroimaging departments, community sources (general practices, outpatient clinics, rehabilitation units) and death certificates^25^. Suspected cases are screened according to World Health Organization ICD-10 diagnostic criteria throughout the study period to maintain consistency^26,27^. All diagnoses were confirmed by study clinicians via review of medical records, neuroimaging, and laboratory results, as previously described in detail^25^. Capture-recapture analyses have previously demonstrated case ascertainment of 88%^28^. An updated internal analysis suggests this remains consistent.

### Study Population, Exposure and Outcomes

Participants included all residents aged ≥18 years with first-ever stroke registered in the SLSR between January 1, 1995, and December 31, 2024. Ethnicity was self-reported at recruitment using UK Office for National Statistics (ONS) levels 1 and 2 census categories^29^. We retained Black African and Black Caribbean as separate subcategories (ONS level 2, i.e. subdivisions of the broader “Black” category at level 1), reflecting the area’s ethnic composition, these groups’ distinct migration histories and previous studies ^10^. Sufficient sample sizes enabled separate analysis of these groups. Asian, Mixed-ethnic groups, and other non-White groups were combined into a single category “other minority ethnic groups” (hereafter Other) to ensure stable estimates. Final categories were: White, Black African, Black Caribbean, and Other.

The primary outcome was first-ever stroke incidence calculated as the number of new stroke cases per 100,000 person-years. Incidence was first calculated for all stroke subtypes combined (hereafter “All Strokes”), then separately by subtype: ischemic stroke, primary intracerebral haemorrhage (PICH), and subarachnoid haemorrhage (SAH), based on imaging and clinical assessment.

### Covariates

The Index of Multiple Deprivation (IMD) was the indicator of SES, a composite measure of relative deprivation at the small-area level (lower super output area [LSOA] comprising ∼1,500 residents)^30^, combining seven weighted domains: income, employment, education, health, crime, barriers to housing and services and living environment^30^. All English LSOAs are ranked and grouped into national quintiles (quintile-1 = most deprived; quintile-5 = least deprived). IMD scores are periodically updated (first in 2004, then 2007, 2010, 2015, 2019). Each patient’s LSOA stroke onset was matched to the most recent IMD available; those with first stroke before 2004 were assigned 2004 values. Consistent with our previous studies^10^, IMD quintiles 3–5 were combined due to small numbers. Quintiles, rather than raw IMD scores, were used as IMD is a relative measure and absolute scores are not comparable across time^30^.

### Population Denominators

Population denominators were derived from UK census data (1991, 2001, 2011, 2021), for adults (aged ≥18) in the SLSR catchment area, disaggregated by sex, age group (five-year bands matching the European Standard Population), and ethnicity.

IMD quintiles were assigned as follows: 1991 and 2001 electoral wards were mapped to 2001 LSOAs using ONS lookups and assigned 2004 IMD quintiles; 2011 wards were mapped to 2010 LSOAs and assigned 2010 IMD quintiles; 2021 used ward-level 2019 IMD scores directly. Where wards spanned multiple LSOAs, we used mean IMD ranks before assigning quintiles. To ensure geographic consistency over time, we defined a fixed set of LSOAs representing the SLSR catchment and applied this across all years, accommodating ward and borough boundary changes (including 2018–2022 Southwark revisions) by mapping to this constant LSOA frame.

A complete grid of all combinations (year, sex, categorical age group, ethnicity, IMD quintile) was constructed. Population estimates for inter-census years (1992–2000, 2002–2010, 2012–2020) and post-census years (2022–2024) were derived by linear interpolation rom census data within this fixed LSOA set.

### Statistical Analysis

Analyses were conducted using a complete case approach, excluding observations with missing data for ethnicity, socioeconomic status, or stroke subtype. The proportion of missing data for these variables was low (<5%), and no imputation was performed. Crude incidence rates were as first-ever strokes divided by person-years at risk, estimated overall and by ethnicity, sex, and subtype. Standardised rates used direct standardisation with the 2013 European Standard Population^31^, with 95% confidence intervals (CIs) from the Fay–Feuer gamma method to account for small event number for some stroke subtypes^32^. Temporal trends used Poisson regression at the stratum level (period × age group × sex), with stroke counts as outcome, log person-years as offset, and age group and sex as covariates. Incidence rate ratios (IRRs) compared each period with the previous one.

Ethnic inequalities in incidence were assessed using Poisson regression with ethnicity, period, age group, and sex as covariates and stroke counts as outcome. IRRs compared each ethnic group with the White population, adjusted first for age and sex, then additionally for IMD. To assess changes in over time, we fitted Poisson models comparing periods in between them. Temporal changes were expressed as the ratio of IRRs between consecutive periods, representing the relative change in ethnic differences in stroke incidence.

To examine whether ethnicity-incidence association varied by IMD, we fitted Poisson models with interaction terms between ethnicity and IMD variables, adjusting for age and sex. Marginal incidence rates for each ethnic-SES subgroup were derived using the *emmeans*^33^, weighted by the observed age-sex distribution of person-years to prevent bias from demographic differences. Estimates were expressed per 100,000 person-years with 95% CIs from 1,000 bootstrap simulations.

Model assumptions (linearity and overdispersion) were assessed using residual plots, deviance statistics, and goodness-of-fit tests. Analyses used R version 4.4.2, with *dplyr* and *tidyr*, *ggplot2*, *lmtest*, *emmeans*, and *writexl* packages

## Results

### Study population

Between 1995–2024, 7,726 people with first-ever stroke were registered in the SLSR (Table 1 and Supplementary Table 1). The catchment area’s ethnic composition changed substantially: White residents declined from 76% in the 1991 census to 60% in the 2021 census, while Black African residents increased from 6% to 13%, and Black Caribbean decreased from 9% to 7% (Supplementary Figure 1). The population aged ≥65 years declined from 17% to 9% (p<0.001), with White and Black Caribbean populations maintaining higher proportions of older adults throughout (Supplementary Table 2). The proportion of the population living in lowest IMD quintile areas fell from 66% to 18% (p<0.001).

**Table 1.**
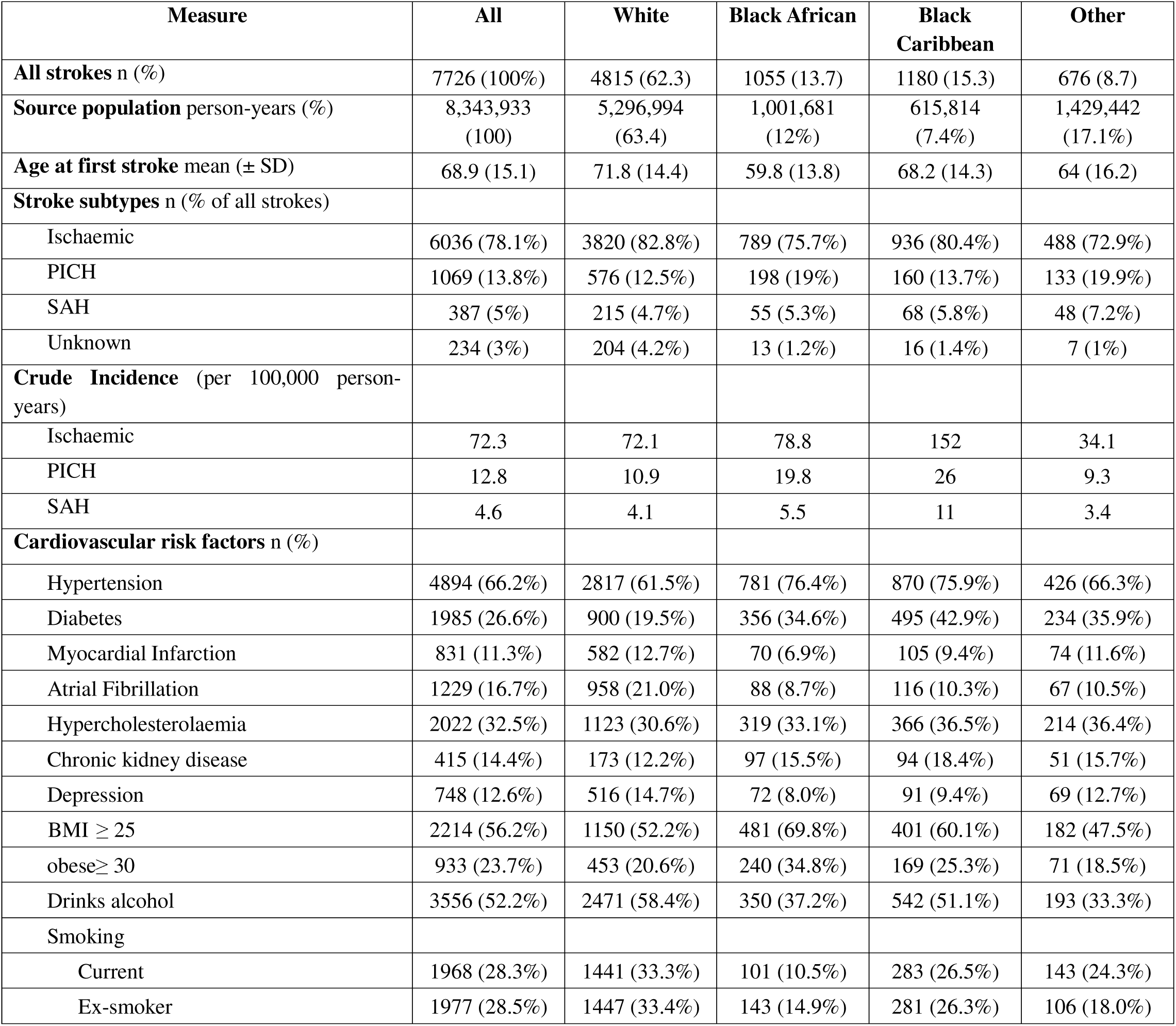
Summary of Stroke Incidence, Subtype Distribution, and Crude Incidence by Ethnic Group (ICD-10 Defined Cases, SLSR, 1995-2024)

### Overall trends in stroke incidence

Standardised stroke incidence decreased by 28% from 1995–1999 to 2005–2009 (IRR = 0.81 [0.75–0.88]), stabilised through 2010–2019, then increased by 17% in 2020–2024 (IRR 019=1.20 [1.11–1.29]; Table 2). Similar patterns were observed for ischaemic stroke (18% reduction then 11% increase) and PICH (41% reduction then 50% increase). SAH trends were less consistent due to small numbers (Supplementary Table 3).

**Table 2.**
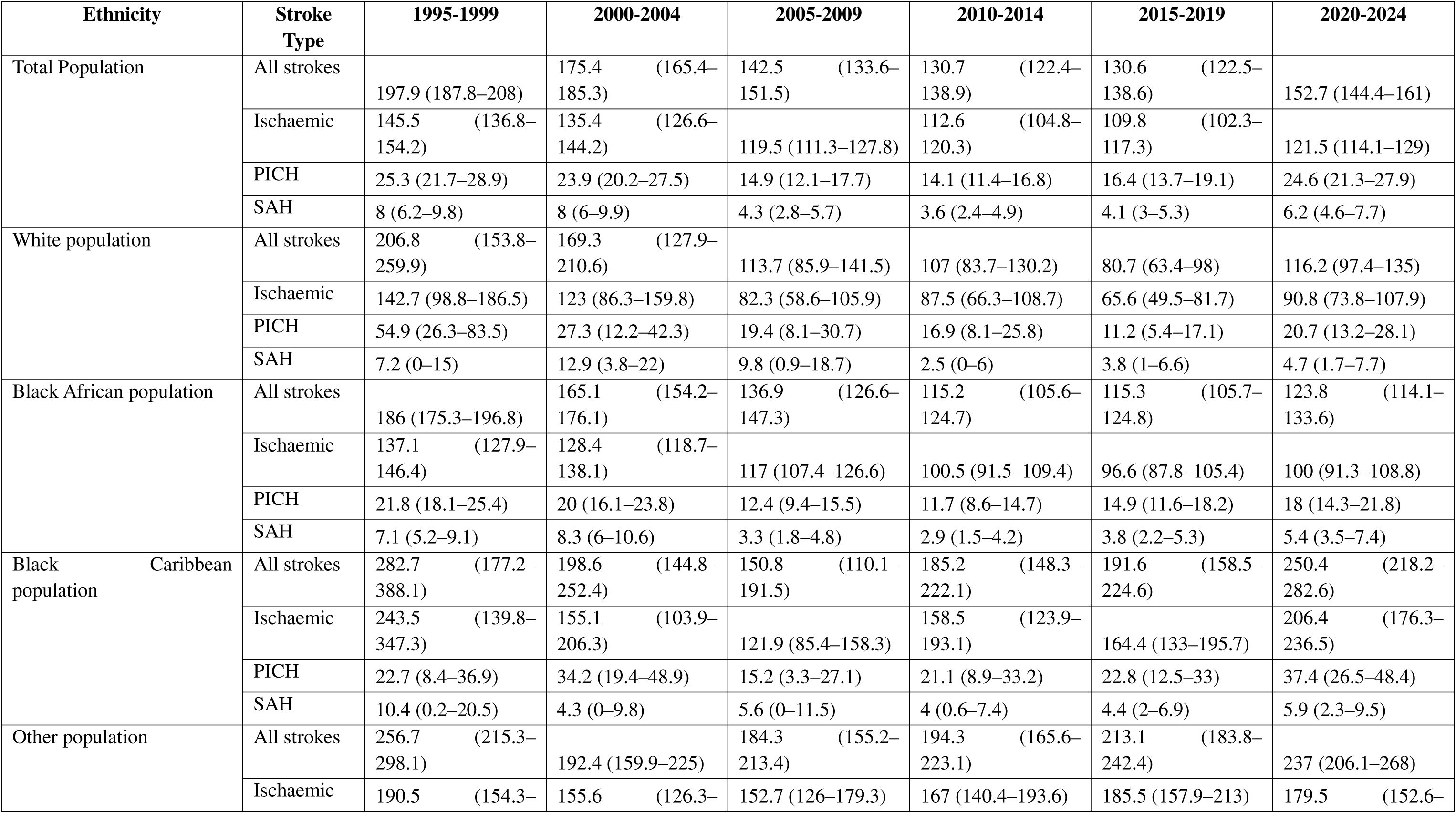
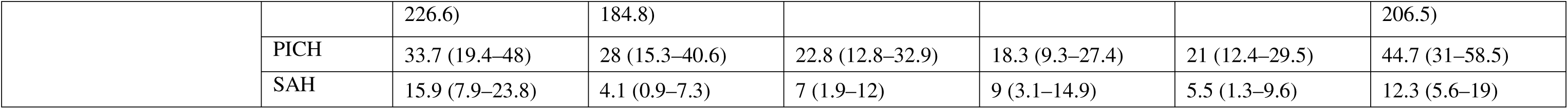
Age- and sex-standardised incidence rates (per 100,000 person-years) with 95% confidence intervals for first-ever stroke by ethnic group, stroke subtype, and time period, South London Stroke Register, 1995–2024.

### Ethnic inequalities in stroke incidence over time

Throughout the 30-year period, standardised stroke incidence remained consistently higher in Black African and Black Caribbean populations than in the White population (Figure 1). In 1995–1999, IRR were 1.57 (95% CI 1.23–2.00) for Black African and 1.52 (1.31–1.77) for Black Caribbean populations compared to the White group (Table 3).

**Figure 1.**
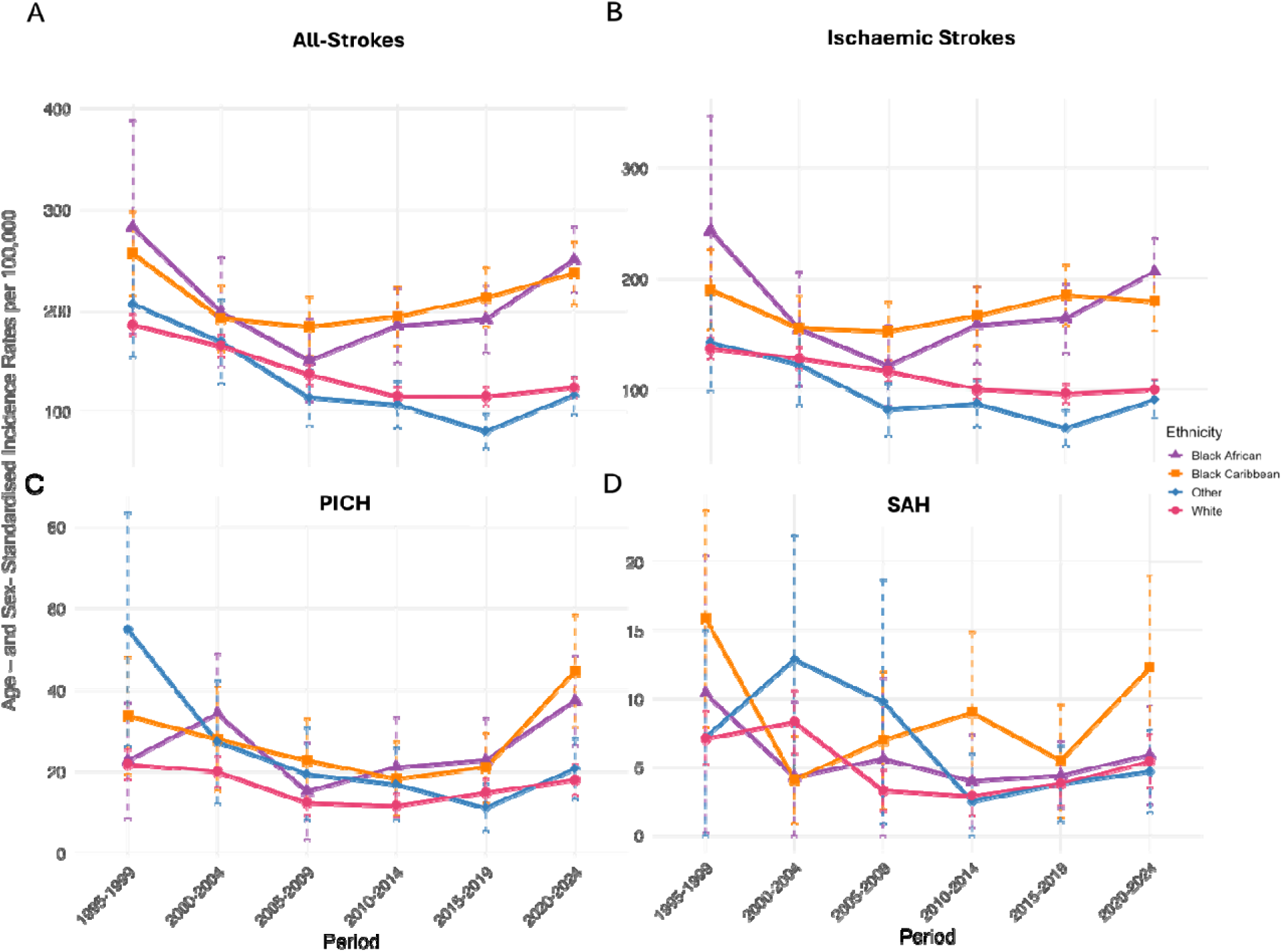
Age- and Sex-Standardised Stroke Incidence Rates by Ethnicity and Stroke Subtype for the total population. 1995-2024 Age and sex-standardised incidence rates for total and subtype strokes per 100,000 population across time periods, stratified by ethnicity. Rates are standardised to the 2013 ESP. Panel A shows incidence for all-strokes while subtypes shown in panels B-D include Ischaemic, PICH, and SAH, with corresponding 95% co intervals.

**Table 3.**
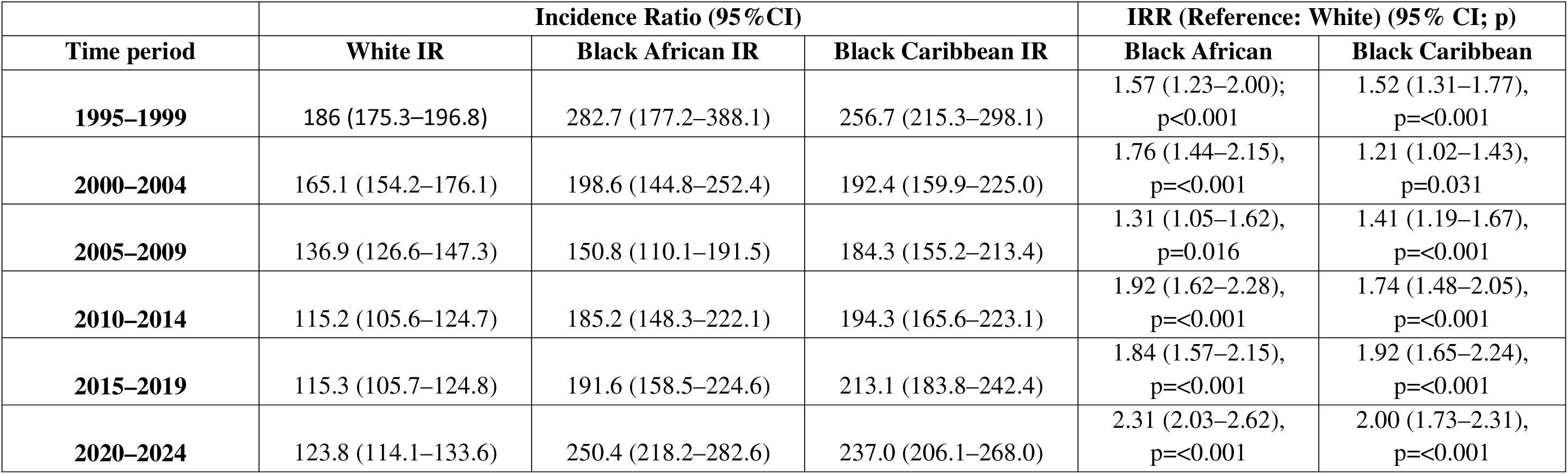
Age-, Sex-, and SES-Adjusted Stroke Incidence and IRRs by Ethnic Group, South London Stroke Register, 1995–2024.

By 2020–2024, standardised all-stroke incidence was 2.31-fold higher (95% CI 2.03–2.62) in Black African and 2-fold higher (1.73–2.31) in Black Caribbean populations compared with White populations. Comparing 2020–2024 to 2015–2019, inequalities for Black African populations widened significantly (IRR 1.30, 1.10–1.60; p=0.01), whilst inequalities for Black Caribbean populations showed no further widening (IRR 1.03, 0.83–1.27; Table 4). This inequality arises from diverging trends: initial reduction over time followed by stability in the White population; initial decline followed by increases for the Black African population; and decline then plateau for the Black Caribbean population (Supplementary Table 4).

**Table 4.** Change in Ethnic Inequalities in Stroke Incidence Over Time (Ratio of Incidence Rate Ratios, Age-and Sex-Adjusted)

| **Ethnic group (vs White)** | **Period comparison** | **Ratio-of-IRR (95% CI)** | **p-value** | **Direction** |
| --- | --- | --- | --- | --- |
| **White (Reference)** | All comparisons | 1.00 (–) | – | – |
| **Black African** | 1995–1999 → 2000–2004 | 1.23 (0.90–1.68) | 0.19 | Widened |
|  | 2000–2004 → 2005–2009 | 0.74 (0.55–0.99) | 0.04 | Narrowed |
|  | 2005–2009 → 2010–2014 | 1.47 (1.12–1.93) | 0.005 | Widened |
|  | 2010–2014 → 2015–2019 | 0.94 (0.75–1.18) | 0.60 | Narrowed |
|  | 2015–2019 → 2020–2024 | 1.30 (1.06–1.58) | 0.01 | Widened |
| **Black Caribbean** | 1995–1999 → 2000–2004 | 0.82 (0.65–1.03) | 0.09 | Narrowed |
|  | 2000–2004 → 2005–2009 | 1.18 (0.93–1.51) | 0.18 | Widened |
|  | 2005–2009 → 2010–2014 | 1.18 (0.93–1.50) | 0.16 | Widened |
|  | 2010–2014 → 2015–2019 | 1.16 (0.92–1.45) | 0.20 | Widened |
|  | 2015–2019 → 2020–2024 | 1.03 (0.83–1.27) | 0.81 | Widened |
| **Other ethnic groups** | 1995–1999 → 2000–2004 | 1.04 (0.76–1.42) | 0.82 | Widened |
|  | 2000–2004 → 2005–2009 | 0.77 (0.56–1.05) | 0.10 | Narrowed |
|  | 2005–2009 → 2010–2014 | 0.98 (0.72–1.32) | 0.88 | Narrowed |
|  | 2010–2014 → 2015–2019 | 0.87 (0.65–1.17) | 0.36 | Narrowed |
|  | 2015–2019 → 2020–2024 | 1.41 (1.09–1.83) | 0.009 | Widened |

The absolute difference in standardised incidence between Black African and White populations increased from 84.8 per 100,000 in 1995–1999 to 97.7 per 100,000 in 2020–2024. In 1995–1999, mean age at first stroke at first stroke was 58.6 ± 15.3 years in Black African (p<0.001 vs White) and 65.3 ± 13.5 years in Black Caribbean (p<0.001 vs White) populations, compared with 73.6 ± 12.8 years in White groups. By 2020–2024, this age-gap narrowed to 9 years for Black African (Mean ± SD Black African vs White = 61.6 ± 13.4 vs 70.1 ± 14.8 years, p<0.001) and was no longer significant for Black Caribbean populations (Mean ± SD Black Caribbean vs White = 69.5 ± 13.9 vs 70.1 ± 14.8 years, p=1.00).

These ethnic inequalities were consistent across stroke subtypes (Supplementary Table 5). In 1995–1999, ischaemic stroke incidence was 1.60-fold higher (95% CI 1.19–2.15) in Black African and 1.46-fold higher (1.22–1.75) in Black Caribbean populations compared with White groups. By 2020–2024, these inequalities increased, with IRRs of 2.22 (1.93–2.57) and 1.86 (1.58–2.19), respectively. For PICH, incidence was 2.18-fold (1.24–3.82) and 1.83-fold (1.23–2.74) higher in 1995–1999, increasing to 3.04 (2.24–4.12) and 2.59 (1.83–3.66) by 2020–2024 for Black African and Black Caribbean groups, respectively. SAH rates were lower overall and differences less consistent, though remained elevated for Black Caribbean populations (IRR 2.61, 1.47–4.63 in 2020–2024).

### Role of SES in ethnic inequalities

Adjustment for IMD attenuated but did not eliminate the ethnic inequalities (Table 5 and Supplementary Table 6). In 2020–2024, IRRs compared with White populations reduced from 2.31 unadjusted (95% CI 2.03–2.62) to 2.03 adjusted (1.78–2.32) for Black African populations, and from 2.00 (1.73–2.31) to 1.79 (1.54–2.07) for Black Caribbean populations.

**Table 5.** Age-, sex-, and IMD-adjusted incidence rate ratios with 95% confidence intervals and p-values for first-ever stroke by ethnic group, stroke subtype, and time period in the total population, South London Stroke Register, 1995–2024

| period | ethnicity | IRR (95%CI); p value |
| --- | --- | --- |
| 1995-1999 | White | Ref. |
|  | Black African | 1.67 (1.31–2.14), p=<0.001 |
|  | Black Caribbean | 1.65 (1.41–1.93), p=<0.001 |
|  | Other | 1.10 (0.87–1.38), p=0.432 |
| 2000-2004 | White | Ref. |
|  | Black African | 1.85 (1.51–2.27), p=<0.001 |
|  | Black Caribbean | 1.26 (1.06–1.51), p=0.010 |
|  | Other | 1.08 (0.87–1.35), p=0.481 |
| 2005-2009 | White | Ref. |
|  | Black African | 1.27 (1.02–1.58), p=0.032 |
|  | Black Caribbean | 1.31 (1.10–1.55), p=0.002 |
|  | Other | 0.90 (0.72–1.12), p=0.335 |
| 2010-2014 | White | Ref. |
|  | Black African | 1.71 (1.44–2.03), p=<0.001 |
|  | Black Caribbean | 1.54 (1.31–1.82), p=<0.001 |
|  | Other | 0.81 (0.65–0.99), p=0.042 |
| 2015-2019 | White | Ref. |
|  | Black African | 1.60 (1.37–1.87), p=<0.001 |
|  | Black Caribbean | 1.78 (1.52–2.08), p=<0.001 |
|  | Other | 0.69 (0.56–0.84), p=<0.001 |
| 2020-2024 | White | Ref. |
|  | Black African | 2.03 (1.78–2.32), p=<0.001 |
|  | Black Caribbean | 1.79 (1.54–2.07), p=<0.001 |
|  | Other | 0.96 (0.82–1.13), p=0.654 |

Marginal incidence analysis showed clear socioeconomic gradients across all ethnic groups, with steeper differences among Black populations.

Marginal incidence analysis and tests for interaction between ethnicity and SES showed strong SES gradients, within all ethnic groups, with steeper gradients observed among Black populations (Figure 2; Supplementary Table 7-8). In the most deprived quintile (IMD 1), stroke incidence was highest among Black African (191.4, 95% CI 176.0–209.0;) and Black Caribbean populations (152.2, 139.8–165.0). The IMD 1–3/5 difference was ∼110 for Black African versus ∼38 for White. Supplementary Table 8 shows corresponding IRRs for IMD–ethnicity interactions.

**Figure 2.**
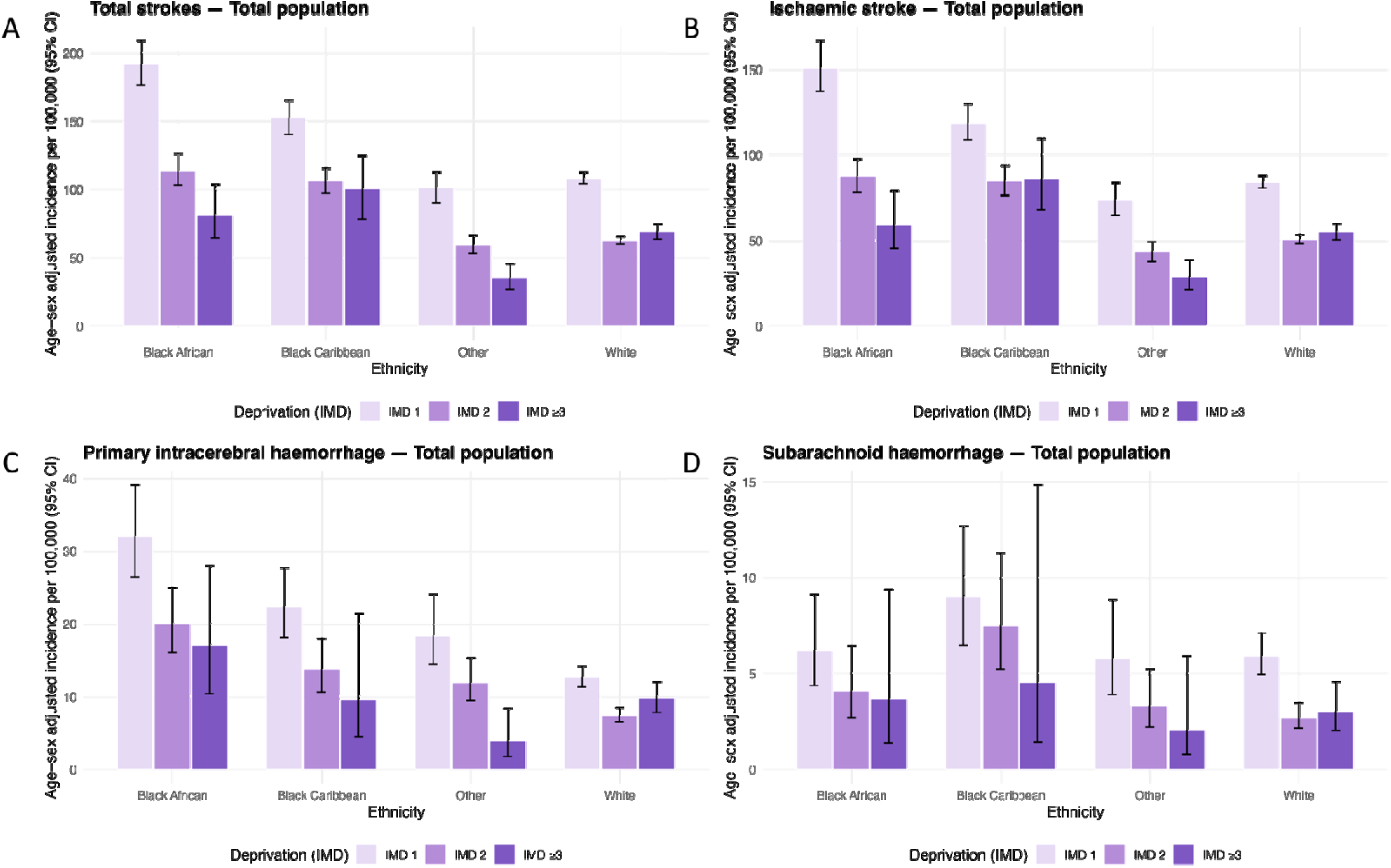
Age- and sex-adjusted marginal incidence rates (per 100,000 person-years) with 95% confidence intervals for first-ever stroke by ethnic group and IMD quintile, South London Stroke Register, 1995–2024. Age and Sex-standardised incidence rates (per 100,000 person-years) for (A) total stroke, (B) ischaemic stroke, (C) PICH, and (D) SAH in the total population, stratified by ethnicity and IMD quintile. IMD quintile 1 represents the most deprived areas, while quintiles 3–5 were combined due to small case counts in less deprived strata for some ethnic groups. Error bars represent 95% CI. Detailed estimates are provided in Supplementary Table 8.

### Impact of sex differences

Ethnic inequalities were observed in both sexes with generally similar patterns (Supplementary Table 4-8). The highest relative risks were seen in Black African men in the most deprived areas: for all strokes (IRR 3.10, 2.64–3.64), ischaemic stroke (3.02, 2.52–3.63), and PICH (3.64, 2.48–5.33). For SAH, the highest IRR was observed in Black Caribbean women (IRR 3.24, 95% CI 1.54–6.82).

## Discussion

This study examined standardised stroke incidence over three decades in South London, overall and by ethnicity, including ethnic inequalities over time, and the role of SES. Between 1995 and 2024, all stroke incidence declined for the total population by 28% from 198 to 143 per 100,000 (1995–1999 to 2010–2014) before increasing by 17% to 167 per 100,000 in 2020–2024. This recent increase was primarily observed in Black African and Black Caribbean populations, while rates in White populations remained stable. Ethnic inequalities persisted and lately widened, with Black African and Black Caribbean people experiencing higher incidence and younger onset, particularly for haemorrhagic subtypes, partly reflecting the population’s younger age. Adjustment for SES did not eliminate these inequalities, though clear socioeconomic gradients were evident and steepest among minority groups. The highest rates of ischaemic stroke and PICH occurred in the in the most deprived Black African populations and of SAH in the most deprived Black Caribbean populations.

Our findings align with US population-based studies^4,8,34^. The Northern Manhattan^4^ and Greater Cincinnati/Northern Kentucky studies both reported persistently higher stroke incidence among Black compared with White populations^8^. Our study extends this evidence in three ways: First, we quantified how ethnic inequalities evolved over three decades, using consistent methods and complete case ascertainment in a multi-ethnic UK population. Second, we demonstrate that inequalities not only persisted but widened over time. Third, by incorporating SES gradients through interaction analyses, we show that ethnicity and deprivation act jointly rather than independently, with groups facing both disadvantages experiencing the highest stroke incidence; reported for the first time in the UK.

Our previous SLSR research showed higher prevalence of hypertension, diabetes, and obesity among Black African and Black Caribbean people at the time of first stroke. Socioeconomic patterns, however, differed: the Black Caribbean population had lower SES, whereas the Black African population often had higher education and non-manual occupations despite residing in deprived neighbourhoods^10^. Mediation analyses have shown that traditional cardiovascular risk factors, particularly hypertension and diabetes, explain at least 30% of ethnic inequalities in stroke incidence^35,36^. In the NHANES Epidemiologic Follow-up Study, these factors accounted for about one-third of the Black–White inequality in stroke risk^35^, while in the REGARDS cohort, nearly 40% was explained by atrial fibrillation, diabetes, hypertension, heart disease, smoking, left ventricular hypertrophy, and antihypertensive use^36^. In REGARDS, hypertension alone accounted for half, and diabetes for one-third of the total mediation, highlighting these two conditions as the strongest causal pathways between ethnicity and stroke incidence^36^.

Despite decades of recognition, hypertension and diabetes remain inequitably distributed and controlled^13,14,37^. While global frameworks by the European Stroke Organisation and World Stroke Organisation have prioritised prevention, the guidelines remain largely generic, with limited focus on ensuring equitable uptake and adapting interventions to the needs of high-risk or deprived populations^38,39^. In the UK, national strategies including the Fit for the Future: 10-Year Health Plan and the Core20PLUS5 framework emphasise prevention and early detection of cardiovascular risk factors, particularly targeting the most deprived 20% of the population^40,41^. The NHS Health Checks programme exemplifies this approach, offering cardiovascular risk screening to adults aged 40–74 for primary prevention^42^. However, achieving equitable benefit from these programmes requires understanding and addressing barriers to uptake. Evidence shows that uptake remains low^43^, with limited effectiveness in translating identified risks into preventive treatment for high-risk individuals^44^, particularly among deprived and ethnic minority groups^43^. In south London, programmes such as *Vital 5*, the *Cardiovascular Disease (CVD) Inequalities Scheme*^45,46^, and *Clinical Effectiveness South East London (CESEL)*^47,48^ aim to address hypertension control inequalities through community and primary care partnerships, though their impact on clinical outcomes such as stroke remains be evaluated.

An important limitation of current prevention efforts is their timing. Our findings show disproportionately earlier stroke incidence in deprived and minority ethnic groups, with Black African populations experiencing first stroke an average of 12 years younger than White peers. Although this partly reflects the younger age structure of the source populations, age-standardised incidence rates confirm that these represent genuine inequalities rather than demographic artefacts, suggesting that CVD risk detection and intervention is happening too late: NHS Health Checks in the UK start at age 40. Enabling health checks earlier in high-risk populations^49^ would enable early detection and control of hypertension and diabetes before stroke risk accumulates. Implementing earlier screening itself is a challenge, as current cardiovascular risk prediction tools are inadequate for this purpose in young adults^50,51^. The risk estimates from these tools are heavily weighted by age, and systematically underestimate lifetime risk in younger adults with lifelong exposure to disadvantage and multiple risk factors^42^. In younger people with elevated risk factors, we lack risk prediction models which usefully discriminate between higher and lower CVD risk: hampering our ability to make treatment decisions^52–54^. There is a clear need for risk prediction tools which adopt a life-course approach, capturing cumulative exposure.

Multiple factors perpetuate poor hypertension control in ethnic minority populations, including mistrust in healthcare services^55^, inadequate treatment intensification during follow-up^14,56^, and differential prescribing practices by ethnicity^14,57^. The COVID-19 pandemic further compounded these barriers, with stroke incidence nationally declining sharply between March and July 2020, probably because health services had reduced capacity and only people with severe illness sought care^58^. Subsequently, national post-diagnosis stroke rates increased by around 13% (June 2023–May 2024 vs January–February 2020), with marked variation by geographic region and SES^58^. In addition, primary care consultations, risk-factor monitoring, and prescribing declined acutely and disproportionately among Black and deprived populations^59^. Subsequently, national post-diagnosis stroke rates increased by around 13% (June 2023–May 2024 vs January–February 2020), with marked variation by geography and deprivation^58^. Nevertless, these inequalities preceded the pandemic; evidence shows that in the UK, Black patients were consistently less likely to achieve target blood pressure control or receive timely treatment intensification^14^. This challenges the overemphasis on biological explanations for the higher cardiovascular burden in Black ethnic groups and highlights the necessity of understanding ethnicity within the structural and socioeconomic contexts in which they are experienced^60^.

Structural and environmental determinants also contribute to the unequal incidence of stroke. In London, Black African and Caribbean populations are more likely to experience overcrowded or poor-quality housing and insecure tenure, which directly contribute to area-level deprivation^61^. These conditions reflect structural discrimination and residential segregation, exacerbating cardiovascular risk through chronic stress, poor air quality, and heat vulnerability^62^. Ethnic minority children in deprived areas are disproportionately exposed to air pollutants such as PM□.□, PM□□, and NO□, known to increase hypertension, diabetes, and overall cardiovascular risk^63,64^. These exposures create a form of “double jeopardy,” where ethnic minority groups facing socioeconomic deprivation experience compounded risks that amplify inequalities and contribute to premature stroke incidence^65^.

Strengths of this study include a population-based registry with 30 years of prospective data collection, active multi-source case ascertainment, and linkage to census denominators, enabling robust age- and sex-standardised stroke incidence rates. The ethnic diversity of South London allowed distinction between Black African and Black Caribbean groups, typically combined despite different migration histories, revealing important heterogeneity within Black populations.

Ethnicity was self-reported and grouped into broad categories, which may mask differences in heterogeneous groups. SES was measured using area-level deprivation, and IMD quintiles were collapsed; limiting individual-level interpretation as IMD reflects neighbourhood rather than personal circumstances. Earlier study periods used IMD 2004 scores retrospectively, which may not fully capture temporal changes in deprivation, and changes in IMD over time may reflect postcode reclassification and local regeneration rather than true socioeconomic improvement. A further limitation is the small number of SAH events, leading to wide confidence intervals.

Future research should prioritise how to measure, detect, and manage cardiovascular risk in equitable ways that effectively reduce the inequalities mediating unfair differences in stroke incidence. This includes developing tools to better stratify risk and deliver equitable care, as well as investigating how early-life exposures, cumulative disadvantage, and intergenerational factors shape ethnic inequalities across the life course. Prospective cohorts linking childhood deprivation, migration history, and cardiovascular trajectories could clarify causal pathways and identify windows for intervention. Evaluations of targeted prevention programmes, should assess their impact on stroke and cardiovascular outcomes, disaggregated by ethnicity and SES. Replication in national datasets and use of advanced methods, including causal frameworks, would strengthen representativeness and inference on the pathways perpetuating these inequalities. Qualitative work exploring perceptions of cardiovascular risk, healthcare trust, and system barriers should complement quantitative findings. Co-developing interventions with these communities and policymakers will be key to ensuring their effectiveness and sustainability.

## Conclusion

Stroke incidence declined over the past 25 years but has risen again recently, with persisting and widening ethnic inequalities in South London. Black African and Black Caribbean populations, especially deprived areas, continue to experience higher and earlier stroke incidence. The younger age at onset highlights missed prevention opportunities and the urgent need for earlier, more equitable action. Despite international and national initiatives, current prevention frameworks are failing to reach those most at risk. Cardiovascular risk factor detection, diagnosis and management, particularly for hypertension, must be strengthened through proactive case finding and equitable care. New risk prediction models are needed to guide preventive action in younger. Systematic evaluation of prevention strategies are needed to ensure earlier and fairer action and further research using causal modelling is needed to identify the perpetuators of these inequalities and enable their correction in the future.

## Data Availability

All data produced in the present study are available upon reasonable request to the authors.
Data types: Deidentified participant data, Data dictionary
How to access data: Because of the sensitive nature of the data collected for this study,
requests to access the data set for academic use should be made to the South London Stroke
Register (SLSR) team:
https://www.kcl.ac.uk/lsm/research/divisions/hscr/research/groups/stroke/index.aspx

## Acknowledgments

We thank Dr Joseph Mayhew and Dr Sian Howell for their insightful feedback on the policy translation and health system implications of our findings. Their reflections on national and local initiatives; including Core20PLUS5, the Vital 5 programme, and the South East London CVD Inequalities Scheme helped strengthen the discussion on prevention, early detection, and equitable implementation within NHS frameworks.

