## Supplementary Material for "Widening Ethnic Inequalities in Stroke Incidence: A 30-Year Population-Based Analysis of the South London Stroke Register"

**Supplementary Table and Figures Index**

**Supplementary Table 1.** Crude incidence rates (per 100,000 person-years) and mean age at first stroke (with standard deviation) by ethnic group, stroke subtype, and time period in the South London Stroke Register, 1995–2024

|  |  | **1995-1999** | | **2000-2004** | | **2005-2009** | | **2010-2014** | | **2015-2019** | | **2020-2024** | |
| --- | --- | --- | --- | --- | --- | --- | --- | --- | --- | --- | --- | --- | --- |
| **Ethnicity** | **Subtype** | **Crude IR** | **Mean age of stroke (SD)** | **Crude IR** | **Mean age of stroke (SD)** | **Crude IR** | **Mean age of stroke (SD)** | **Crude IR** | **Mean age of stroke (SD)** | **Crude IR** | **Mean age of stroke (SD)** | **Crude IR** | **Mean age of stroke (SD)** |
| Total Population | All-strokes | 139.0 | 71.4 (13.7) | 108.1 | 69.8 (14.8) | 80.4 | 69.5 (15.3) | 73.2 | 67.8 (16.0) | 74.1 | 67.7 (16.0) | 88.8 | 67.0 (15.0) |
|  | Ischaemic | 100.4 | 72.4 (12.1) | 81.2 | 72.0 (13.2) | 65.9 | 70.4 (14.6) | 61.6 | 69.1 (15.5) | 59.6 | 69.4 (15.4) | 69.8 | 68.0 (14.6) |
|  | PICH | 18.0 | 68.8 (14.6) | 15.4 | 64.6 (15.8) | 9.3 | 67.0 (17.3) | 8.5 | 63.0 (15.6) | 10.9 | 63.1 (16.7) | 14.6 | 65.3 (15.5) |
|  | SAH | 7.7 | 53.0 (17.8) | 6.7 | 52.6 (15.8) | 3.1 | 57.5 (17.0) | 2.8 | 54.3 (17.2) | 3.6 | 53.6 (13.6) | 4.2 | 56.8 (15.1) |
| White | All-strokes | 152.2 | 73.6 (12.8) | 116.4 | 72.5 (14.0) | 85.4 | 72.1 (14.8) | 68.0 | 70.5 (15.6) | 64.8 | 70.2 (15.5) | 67.2 | 70.1 (14.8) |
|  | Ischaemic | 111.3 | 73.9 (11.8) | 89.6 | 74.2 (12.7) | 71.9 | 72.7 (14.2) | 58.5 | 71.5 (15.0) | 52.9 | 71.1 (15.2) | 54.7 | 70.5 (14.5) |
|  | PICH | 17.8 | 72.0 (13.6) | 13.6 | 70.6 (13.2) | 8.3 | 72.9 (15.2) | 7.2 | 66.3 (16.7) | 9.2 | 68.3 (16.5) | 9.2 | 71.6 (16.0) |
|  | SAH | 6.6 | 57.0 (16.6) | 7.2 | 53.8 (16.6) | 2.7 | 55.7 (18.1) | 2.2 | 57.0 (17.9) | 2.7 | 59.1 (12.3) | 3.2 | 60.0 (14.2) |
| Black African | All-strokes | 64.5 | 58.6 (15.3) | 79.4 | 55.4 (12.8) | 60.5 | 58.5 (13.6) | 96.7 | 58.8 (13.3) | 110.0 | 60.4 (14.5) | 168.6 | 61.6 (13.4) |
|  | Ischaemic | 43.3 | 62.3 (14.9) | 49.4 | 58.9 (12.3) | 46.7 | 60.1 (13.1) | 75.9 | 60.8 (12.9) | 84.5 | 63.1 (14.1) | 129.5 | 63.3 (12.8) |
|  | PICH | 12.0 | 53.1 (9.3) | 23.6 | 49.6 (11.0) | 9.5 | 50.2 (13.9) | 15.2 | 52.3 (11.5) | 19.0 | 54.1 (12.1) | 32.3 | 58.1 (11.9) |
|  | SAH | 6.4 | 43.4 (18.9) | 3.6 | 45.9 (15.1) | 3.2 | 54.9 (11.5) | 4.5 | 46.8 (14.7) | 6.5 | 43.8 (8.7) | 6.0 | 45.0 (14.3) |
| Black Caribbean | All-strokes | 191.4 | 65.3 (13.5) | 154.7 | 67.6 (12.7) | 167.7 | 67.5 (13.4) | 189.7 | 68.2 (15.7) | 205.1 | 70.2 (15.4) | 218.9 | 69.5 (13.9) |
|  | Ischaemic | 134.6 | 68.5 (9.6) | 123.1 | 69.2 (11.5) | 135.8 | 68.5 (12.8) | 162.3 | 69.3 (15.5) | 174.3 | 71.4 (14.8) | 169.0 | 70.3 (13.8) |
|  | PICH | 27.9 | 60.1 (14.6) | 21.4 | 65.0 (12.8) | 21.6 | 67.1 (13.3) | 17.2 | 65.3 (13.7) | 23.3 | 63.5 (18.0) | 38.5 | 68.5 (12.3) |
|  | SAH | 19.9 | 47.4 (18.0) | 7.1 | 44.0 (8.1) | 8.2 | 53.8 (16.8) | 10.1 | 56.0 (16.9) | 6.5 | 59.7 (11.5) | 11.4 | 60.7 (17.1) |
| Other | All-strokes | 70.9 | 64.7 (13.6) | 60.0 | 63.5 (15.2) | 38.1 | 63.3 (16.5) | 35.8 | 66.0 (17.1) | 35.3 | 63.6 (17.6) | 56.9 | 63.0 (16.3) |
|  | Ischaemic | 47.8 | 65.4 (12.0) | 38.2 | 67.1 (12.6) | 27.0 | 64.5 (16.1) | 28.7 | 67.0 (17.8) | 25.2 | 67.5 (17.0) | 42.3 | 64.4 (16.4) |
|  | PICH | 16.8 | 68.1 (12.7) | 13.0 | 56.2 (19.4) | 7.5 | 57.4 (19.2) | 5.8 | 64.7 (11.2) | 6.6 | 56.7 (15.4) | 11.1 | 59.6 (16.7) |
|  | SAH | 5.3 | 47.3 (20.1) | 6.8 | 55.3 (14.4) | 2.2 | 74.7 (6.2) | 1.4 | 51.6 (20.2) | 3.5 | 48.3 (15.1) | 3.6 | 56.6 (10.6) |

**Supplementary Table 2. A**ge distribution of the adult population (aged ≥18 years) by ethnic group in the South London Stroke Register catchment area, comparing 1991 and 2021 (census-derived)

| **Ethnicity** | **Age group** | **1991 (n)** | **1991 (%)** | **2021 (n)** | **2021 (%)** | **p-value** |
| --- | --- | --- | --- | --- | --- | --- |
| **White** | 18-19 | 3749 | 2.2 | 3708 | 1.9 | <0.001 |
|  | 20-24 | 19340 | 11.6 | 22987 | 11.5 |  |
|  | 25-29 | 27513 | 16.5 | 43336 | 21.7 |  |
|  | 30-34 | 20570 | 12.3 | 31284 | 15.7 |  |
|  | 35-39 | 13787 | 8.3 | 19841 | 9.9 |  |
|  | 40-44 | 11987 | 7.2 | 14463 | 7.2 |  |
|  | 45-49 | 9679 | 5.8 | 12484 | 6.3 |  |
|  | 50-54 | 9031 | 5.4 | 12177 | 6.1 |  |
|  | 55-59 | 9288 | 5.6 | 11422 | 5.7 |  |
|  | 60-64 | 9866 | 5.9 | 8709 | 4.4 |  |
|  | 65-69 | 10064 | 6 | 6111 | 3.1 |  |
|  | 70-74 | 7950 | 4.8 | 5405 | 2.7 |  |
|  | 75-79 | 6955 | 4.2 | 3213 | 1.6 |  |
|  | 80-84 | 4362 | 2.6 | 2267 | 1.1 |  |
|  | 85 | 2952 | 1.8 | 2287 | 1.1 |  |
|  | Total Population | 167093 | 100 | 199694 | 100 |  |
| **Black African** | 18-19 | 505 | 3.8 | 1697 | 3.9 | <0.001 |
|  | 20-24 | 1917 | 14.5 | 4484 | 10.3 |  |
|  | 25-29 | 3270 | 24.7 | 4556 | 10.5 |  |
|  | 30-34 | 2791 | 21.1 | 4348 | 10 |  |
|  | 35-39 | 1609 | 12.2 | 4073 | 9.3 |  |
|  | 40-44 | 1027 | 7.8 | 4255 | 9.8 |  |
|  | 45-49 | 761 | 5.8 | 4430 | 10.2 |  |
|  | 50-54 | 677 | 5.1 | 4953 | 11.4 |  |
|  | 55-59 | 304 | 2.3 | 4208 | 9.7 |  |
|  | 60-64 | 206 | 1.6 | 2963 | 6.8 |  |
|  | 65-69 | 88 | 0.7 | 1507 | 3.5 |  |
|  | 70-74 | 40 | 0.3 | 855 | 2 |  |
|  | 75-79 | 26 | 0.2 | 592 | 1.4 |  |
|  | 80-84 | 9 | 0.1 | 421 | 1 |  |
|  | 85 | 3 | 0 | 241 | 0.6 |  |
|  | Total Population | 13233 | 100 | 43583 | 100 |  |
| **Black Caribbean** | 18-19 | 773 | 3.7 | 803 | 3.5 | <0.001 |
|  | 20-24 | 2725 | 13.2 | 1958 | 8.5 |  |
|  | 25-29 | 3750 | 18.2 | 1965 | 8.6 |  |
|  | 30-34 | 2702 | 13.1 | 2048 | 8.9 |  |
|  | 35-39 | 1559 | 7.6 | 1799 | 7.9 |  |
|  | 40-44 | 982 | 4.8 | 1865 | 8.1 |  |
|  | 45-49 | 1098 | 5.3 | 1801 | 7.9 |  |
|  | 50-54 | 1703 | 8.3 | 2521 | 11 |  |
|  | 55-59 | 1817 | 8.8 | 2680 | 11.7 |  |
|  | 60-64 | 1582 | 7.7 | 1956 | 8.5 |  |
|  | 65-69 | 1006 | 4.9 | 1053 | 4.6 |  |
|  | 70-74 | 541 | 2.6 | 614 | 2.7 |  |
|  | 75-79 | 259 | 1.3 | 579 | 2.5 |  |
|  | 80-84 | 87 | 0.4 | 639 | 2.8 |  |
|  | 85 | 41 | 0.2 | 631 | 2.8 |  |
|  | Total Population | 20625 | 100 | 22912 | 100 |  |
| **Other** | 18-19 | 788 | 5.3 | 2428 | 4.1 | <0.001 |
|  | 20-24 | 2380 | 15.9 | 8542 | 14.4 |  |
|  | 25-29 | 2747 | 18.3 | 9409 | 15.8 |  |
|  | 30-34 | 2288 | 15.3 | 8256 | 13.9 |  |
|  | 35-39 | 1740 | 11.6 | 6613 | 11.1 |  |
|  | 40-44 | 1228 | 8.2 | 5576 | 9.4 |  |
|  | 45-49 | 898 | 6 | 5005 | 8.4 |  |
|  | 50-54 | 938 | 6.3 | 4397 | 7.4 |  |
|  | 55-59 | 754 | 5 | 3211 | 5.4 |  |
|  | 60-64 | 489 | 3.3 | 2111 | 3.5 |  |
|  | 65-69 | 279 | 1.9 | 1527 | 2.6 |  |
|  | 70-74 | 202 | 1.3 | 982 | 1.7 |  |
|  | 75-79 | 138 | 0.9 | 667 | 1.1 |  |
|  | 80-84 | 65 | 0.4 | 503 | 0.8 |  |
|  | 85 | 53 | 0.4 | 284 | 0.5 |  |
|  | Total Population | 14987 | 100 | 59511 | 100 |  |

**Supplementary Table 3.** Age- and sex-standardised incidence rates by sex (per 100,000 person-years) with 95% confidence intervals for first-ever stroke by ethnic group, stroke subtype, and time period, South London Stroke Register, 1995–2024

|  |  | **1995-1999** | | **2000-2004** | | **2005-2009** | | **2010-2014** | | **2015-2019** | | **2020-2024** | |
| --- | --- | --- | --- | --- | --- | --- | --- | --- | --- | --- | --- | --- | --- |
| **Ethnicity** | **Stroke Type** | **Female** | **Male** | **Female** | **Male** | **Female** | **Male** | **Female** | **Male** | **Female** | **Male** | **Female** | **Male** |
| Total population | All-strokes | 87.3 (81.2–93.4) | 110.6 (102.5–118.7) | 77.7 (71.5–83.9) | 97.7 (89.8–105.5) | 62.5 (57–68.1) | 79.8 (72.8–86.9) | 58.1 (52.8–63.3) | 72.6 (66.2–79) | 54.2 (49.3–59) | 76.1 (69.7–82.6) | 65.2 (60.1–70.3) | 87.5 (81–94.1) |
|  | Ischaemic | 63.2 (57.9–68.4) | 82.3 (75.3–89.3) | 61.9 (56.4–67.4) | 73.5 (66.6–80.4) | 50.7 (45.7–55.7) | 68.7 (62.1–75.2) | 49.6 (44.7–54.5) | 63 (57–69) | 44.6 (40.2–49) | 64.9 (58.9–71) | 51.5 (46.9–56.1) | 70 (64.1–76) |
|  | PICH | 9.8 (7.8–11.9) | 15.5 (12.5–18.5) | 8.3 (6.3–10.3) | 15.6 (12.5–18.6) | 7.9 (6–9.9) | 6.9 (5–8.9) | 5.9 (4.3–7.6) | 8.2 (6.1–10.3) | 7.1 (5.4–8.7) | 9.4 (7.3–11.5) | 10 (8–12) | 14.6 (12–17.2) |
|  | SAH | 4.3 (3–5.7) | 3.7 (2.4–4.9) | 3.8 (2.6–5.1) | 4.1 (2.7–5.6) | 2.4 (1.3–3.4) | 1.9 (0.9–2.9) | 2.2 (1.3–3.1) | 1.4 (0.6–2.2) | 2.5 (1.6–3.4) | 1.7 (0.9–2.4) | 3.3 (2.3–4.3) | 2.9 (1.7–4.1) |
| White population | All-strokes | 82.4 (76–88.8) | 103.6 (95–112.2) | 72.8 (66.1–79.5) | 92.3 (83.6–101.1) | 58.8 (52.5–65) | 77.9 (69.7–86.2) | 50.5 (44.5–56.4) | 64.7 (57.2–72.2) | 46.5 (40.8–52.2) | 68.5 (60.8–76.1) | 51.1 (39.2–63) | 65.1 (50.5–79.7) |
|  | Ischaemic | 59.5 (54.1–65) | 77.6 (70.1–85) | 57.8 (51.9–63.7) | 70.6 (62.9–78.2) | 48.6 (42.9–54.3) | 68.2 (60.4–75.9) | 43.1 (37.6–48.6) | 57.4 (50.4–64.5) | 37.5 (32.3–42.6) | 58.8 (51.6–66) | 38.6 (28.2–49) | 52.3 (38.8–65.8) |
|  | PICH | 8.5 (6.4–10.5) | 13.3 (10.2–16.3) | 7.3 (5.2–9.5) | 12.6 (9.4–15.9) | 6.3 (4.3–8.3) | 6.1 (3.9–8.4) | 5.7 (3.7–7.7) | 5.9 (3.7–8.2) | 6.7 (4.6–8.9) | 8.2 (5.7–10.7) | 10.1 (4.8–15.4) | 10.5 (5.3–15.8) |
|  | SAH | 4 (2.5–5.4) | 3.1 (1.8–4.5) | 3.6 (2.2–5) | 4.7 (2.9–6.5) | 2.1 (0.9–3.2) | 1.2 (0.3–2.1) | 1.5 (0.5–2.4) | 1.4 (0.4–2.3) | 2.3 (1.1–3.5) | 1.5 (0.5–2.4) | 2.4 (0.2–4.7) | 2.3 (0.3–4.3) |
| Black African population | All-strokes | 117 (51.5–182.6) | 165.7 (83.1–248.3) | 96.4 (60.3–132.6) | 102.2 (62.3–142) | 57.8 (32.3–83.2) | 93 (61.2–124.8) | 87.4 (61.9–112.9) | 97.8 (71.1–124.6) | 74.3 (56.2–92.4) | 116.8 (89.2–144.5) | 54 (47.8–60.1) | 69.9 (62.3–77.4) |
|  | Ischaemic | 110 (44.8–175.2) | 133.5 (52.8–214.2) | 77.7 (43.3–112.2) | 77.4 (39.5–115.2) | 49.5 (26.3–72.8) | 72.4 (44.3–100.4) | 77.9 (53–102.7) | 80.7 (56.6–104.7) | 66.3 (48.7–84) | 97.6 (71.7–123.4) | 43.8 (38.2–49.3) | 56.3 (49.5–63) |
|  | PICH | 1.6 (0–3.5) | 21.1 (7–35.2) | 15.8 (5.1–26.5) | 18.4 (8.2–28.6) | 7.7 (0–17.9) | 7.5 (1.4–13.7) | 4.4 (0.9–7.9) | 16.7 (5–28.3) | 5.4 (2–8.7) | 17.4 (7.7–27.1) | 7.8 (5.4–10.1) | 10.3 (7.3–13.2) |
|  | SAH | 0.8 (0–1.9) | 9.6 (0–19.7) | 1.6 (0–3.4) | 2.7 (0–7.8) | 0.6 (0–1.4) | 5 (0–10.9) | 3.5 (0.2–6.7) | 0.5 (0–1.4) | 2.6 (0.7–4.4) | 1.9 (0.2–3.5) | 2.1 (1–3.1) | 3.3 (1.7–5) |
| Black Caribbean population | All-strokes | 110 (83.6–136.4) | 146.7 (114.8–178.5) | 81 (61–101) | 111.5 (85.8–137.1) | 79.7 (61.9–97.6) | 104.6 (81.7–127.5) | 82.7 (65.4–100.1) | 111.6 (88.7–134.5) | 88 (71.1–104.8) | 125.1 (101.2–149.1) | 94.8 (77.7–111.8) | 155.6 (128.3–183) |
|  | Ischaemic | 80 (57.3–102.6) | 110.5 (82.3–138.7) | 70.6 (51.8–89.5) | 84.9 (62.5–107.3) | 62.2 (46.3–78.1) | 90.4 (69–111.8) | 72.1 (55.8–88.3) | 94.9 (73.9–116) | 73.4 (57.9–88.9) | 112.1 (89.3–134.9) | 74.7 (59.3–90) | 131.7 (105.8–157.6) |
|  | PICH | 11.3 (4–18.7) | 22.4 (10.1–34.7) | 4.5 (0–9.1) | 23.5 (11.7–35.2) | 12.6 (5.6–19.7) | 10.2 (3.1–17.3) | 5 (0.8–9.2) | 13.3 (5.2–21.4) | 11.9 (5.8–17.9) | 9.1 (3–15.2) | 14.6 (8.1–21.2) | 22.8 (14.1–31.6) |
|  | SAH | 8.2 (2.1–14.3) | 7.7 (2.5–12.8) | 3.1 (0.5–5.6) | 1 (0–3.1) | 3.9 (0.4–7.4) | 3.1 (0–6.8) | 5.6 (1.3–10) | 3.4 (0–7.4) | 2.7 (0–5.4) | 2.7 (0–5.9) | 4.8 (1.4–8.2) | 1.1 (0–2.3) |
| Other population | All-strokes | 94.2 (59.2–129.1) | 112.7 (72.7–152.6) | 55.7 (33.7–77.7) | 113.6 (78.6–148.6) | 67.2 (44.7–89.8) | 46.5 (30.2–62.7) | 49.1 (33.3–64.9) | 57.9 (40.8–75) | 39.9 (28.2–51.6) | 40.8 (28–53.5) | 96.8 (79.4–114.1) | 140.3 (114.6–165.9) |
|  | Ischaemic | 62.5 (33.5–91.5) | 80.2 (47.3–113) | 40.3 (21–59.7) | 82.7 (51.5–114) | 45.9 (27.2–64.7) | 36.3 (22–50.7) | 40.9 (26.4–55.4) | 46.6 (31.1–62.1) | 34 (23–45.1) | 31.6 (19.9–43.2) | 72.4 (57.5–87.3) | 107.1 (84.7–129.5) |
|  | PICH | 24.8 (6.9–42.6) | 30.1 (7.7–52.5) | 7.1 (0–14.8) | 20.2 (7.2–33.1) | 14.3 (4.1–24.5) | 5.1 (0.3–9.9) | 7.6 (1.4–13.9) | 9.3 (3–15.6) | 3.6 (0.6–6.6) | 7.6 (2.7–12.6) | 14.8 (7.7–21.9) | 30 (18.2–41.7) |
|  | SAH | 6.9 (0–14.6) | 0.3 (0–0.9) | 8.3 (1–15.6) | 4.6 (0–10) | 5.9 (0–12.8) | 3.9 (0–9.5) | 0.5 (0–1.2) | 2 (0–5.5) | 2.3 (0–4.6) | 1.6 (0.1–3) | 9.1 (3.9–14.3) | 3.2 (0–7.4) |

**Supplementary Table 4.** Temporal Changes in Age- and Sex-Standardised Incidence of All-Strokes by Ethnicity, 1995-2024

|  |  |  | **IRR (95% CI), p value** | | | |
| --- | --- | --- | --- | --- | --- | --- |
| **Ethnicity** | **Sex** | **Comparison** | **All stroke** | **Ischaemic** | **PICH** | **SAH** |
| Total population | All | 2000-2004 vs 1995-1999 | 0.89 (0.83–0.96), p=0.002 | 0.94 (0.86–1.02), p=0.135 | 0.96 (0.79–1.17), p=0.692 | 0.91 (0.67–1.24), p=0.550 |
|  |  | 2005-2009 vs 2000-2004 | 0.81 (0.75–0.88), p=<0.001 | 0.89 (0.81–0.98), p=0.014 | 0.64 (0.51–0.80), p=<0.001 | 0.47 (0.32–0.68), p=<0.001 |
|  |  | 2010-2014 vs 2005-2009 | 0.95 (0.87–1.03), p=0.192 | 0.97 (0.89–1.07), p=0.561 | 0.95 (0.74–1.22), p=0.699 | 0.93 (0.61–1.44), p=0.753 |
|  |  | 2015-2019 vs 2010-2014 | 0.97 (0.90–1.06), p=0.526 | 0.93 (0.85–1.02), p=0.121 | 1.22 (0.97–1.53), p=0.095 | 1.22 (0.82–1.84), p=0.319 |
|  |  | 2020-2024 vs 2015-2019 | 1.20 (1.11–1.29), p=<0.001 | 1.16 (1.06–1.26), p=<0.001 | 1.38 (1.14–1.67), p=0.001 | 1.27 (0.90–1.79), p=0.173 |
|  | Female | 2000-2004 vs 1995-1999 | 0.90 (0.81–0.99), p=0.038 | 0.99 (0.87–1.11), p=0.821 | 0.87 (0.63–1.18), p=0.370 | 0.88 (0.58–1.33), p=0.531 |
|  |  | 2005-2009 vs 2000-2004 | 0.80 (0.72–0.90), p=<0.001 | 0.84 (0.73–0.95), p=0.007 | 0.91 (0.65–1.27), p=0.568 | 0.52 (0.31–0.85), p=0.010 |
|  |  | 2010-2014 vs 2005-2009 | 0.95 (0.84–1.07), p=0.392 | 0.99 (0.86–1.14), p=0.892 | 0.80 (0.55–1.14), p=0.211 | 1.03 (0.59–1.79), p=0.923 |
|  |  | 2015-2019 vs 2010-2014 | 0.94 (0.84–1.07), p=0.362 | 0.90 (0.79–1.03), p=0.143 | 1.23 (0.87–1.75), p=0.245 | 1.10 (0.66–1.85), p=0.709 |
|  |  | 2020-2024 vs 2015-2019 | 1.22 (1.09–1.37), p=<0.001 | 1.18 (1.04–1.35), p=0.010 | 1.31 (0.98–1.77), p=0.068 | 1.32 (0.85–2.07), p=0.226 |
|  | Male | 2000-2004 vs 1995-1999 | 0.89 (0.80–0.99), p=0.027 | 0.89 (0.79–1.00), p=0.059 | 1.03 (0.79–1.34), p=0.841 | 0.96 (0.61–1.51), p=0.857 |
|  |  | 2005-2009 vs 2000-2004 | 0.82 (0.73–0.91), p=<0.001 | 0.95 (0.83–1.08), p=0.411 | 0.47 (0.34–0.65), p=<0.001 | 0.41 (0.22–0.71), p=0.002 |
|  |  | 2010-2014 vs 2005-2009 | 0.94 (0.84–1.06), p=0.317 | 0.96 (0.84–1.09), p=0.492 | 1.13 (0.80–1.61), p=0.494 | 0.80 (0.40–1.61), p=0.535 |
|  |  | 2015-2019 vs 2010-2014 | 1.00 (0.89–1.12), p=0.991 | 0.95 (0.84–1.08), p=0.453 | 1.21 (0.89–1.65), p=0.229 | 1.44 (0.76–2.80), p=0.271 |
|  |  | 2020-2024 vs 2015-2019 | 1.18 (1.06–1.31), p=0.002 | 1.14 (1.01–1.27), p=0.030 | 1.43 (1.11–1.85), p=0.007 | 1.20 (0.70–2.06), p=0.511 |
| White | All | 2000-2004 vs 1995-1999 | 0.89 (0.82–0.97), p=0.009 | 0.95 (0.86–1.04), p=0.266 | 0.89 (0.70–1.15), p=0.384 | 1.15 (0.79–1.67), p=0.474 |
|  |  | 2005-2009 vs 2000-2004 | 0.83 (0.75–0.91), p=<0.001 | 0.91 (0.81–1.01), p=0.078 | 0.68 (0.50–0.91), p=0.011 | 0.39 (0.23–0.62), p=<0.001 |
|  |  | 2010-2014 vs 2005-2009 | 0.86 (0.78–0.96), p=0.008 | 0.89 (0.79–1.00), p=0.042 | 0.94 (0.67–1.32), p=0.739 | 0.84 (0.46–1.53), p=0.564 |
|  |  | 2015-2019 vs 2010-2014 | 0.97 (0.87–1.08), p=0.574 | 0.92 (0.82–1.04), p=0.186 | 1.29 (0.94–1.79), p=0.118 | 1.21 (0.68–2.20), p=0.514 |
|  |  | 2020-2024 vs 2015-2019 | 1.08 (0.97–1.20), p=0.176 | 1.06 (0.94–1.20), p=0.337 | 1.09 (0.82–1.46), p=0.547 | 1.28 (0.78–2.15), p=0.336 |
|  | Female | 2000-2004 vs 1995-1999 | 0.89 (0.79–1.00), p=0.047 | 0.98 (0.85–1.12), p=0.775 | 0.84 (0.57–1.21), p=0.350 | 0.96 (0.56–1.62), p=0.877 |
|  |  | 2005-2009 vs 2000-2004 | 0.82 (0.71–0.94), p=0.004 | 0.85 (0.73–0.99), p=0.039 | 0.90 (0.59–1.39), p=0.646 | 0.53 (0.27–1.00), p=0.055 |
|  |  | 2010-2014 vs 2005-2009 | 0.86 (0.74–1.01), p=0.060 | 0.88 (0.74–1.05), p=0.154 | 0.95 (0.60–1.51), p=0.826 | 0.77 (0.34–1.70), p=0.521 |
|  |  | 2015-2019 vs 2010-2014 | 0.92 (0.78–1.09), p=0.333 | 0.88 (0.73–1.06), p=0.166 | 1.13 (0.71–1.79), p=0.614 | 1.30 (0.60–2.91), p=0.508 |
|  |  | 2020-2024 vs 2015-2019 | 1.15 (0.98–1.36), p=0.088 | 1.15 (0.96–1.39), p=0.125 | 1.15 (0.75–1.77), p=0.523 | 1.02 (0.50–2.08), p=0.965 |
|  | Male | 2000-2004 vs 1995-1999 | 0.89 (0.79–1.01), p=0.077 | 0.91 (0.78–1.04), p=0.174 | 0.94 (0.67–1.31), p=0.696 | 1.38 (0.81–2.40), p=0.240 |
|  |  | 2005-2009 vs 2000-2004 | 0.83 (0.72–0.95), p=0.007 | 0.96 (0.82–1.12), p=0.588 | 0.51 (0.33–0.78), p=0.002 | 0.27 (0.12–0.54), p=<0.001 |
|  |  | 2010-2014 vs 2005-2009 | 0.86 (0.74–1.00), p=0.051 | 0.88 (0.75–1.03), p=0.122 | 0.94 (0.57–1.54), p=0.795 | 0.94 (0.37–2.41), p=0.900 |
|  |  | 2015-2019 vs 2010-2014 | 1.00 (0.86–1.17), p=0.948 | 0.95 (0.81–1.12), p=0.554 | 1.47 (0.94–2.32), p=0.096 | 1.11 (0.46–2.75), p=0.822 |
|  |  | 2020-2024 vs 2015-2019 | 1.02 (0.88–1.18), p=0.795 | 0.99 (0.85–1.16), p=0.920 | 1.05 (0.71–1.54), p=0.819 | 1.64 (0.79–3.56), p=0.194 |
| Black African | All | 2000-2004 vs 1995-1999 | 1.07 (0.80–1.45), p=0.646 | 0.97 (0.67–1.41), p=0.874 | 1.76 (0.95–3.47), p=0.083 | 0.53 (0.16–1.67), p=0.283 |
|  |  | 2005-2009 vs 2000-2004 | 0.62 (0.47–0.81), p=<0.001 | 0.75 (0.54–1.04), p=0.085 | 0.34 (0.18–0.61), p=<0.001 | 0.82 (0.23–2.95), p=0.748 |
|  |  | 2010-2014 vs 2005-2009 | 1.34 (1.05–1.73), p=0.022 | 1.34 (1.01–1.78), p=0.046 | 1.45 (0.78–2.80), p=0.252 | 1.32 (0.44–4.37), p=0.630 |
|  |  | 2015-2019 vs 2010-2014 | 0.93 (0.76–1.14), p=0.475 | 0.88 (0.70–1.10), p=0.264 | 1.10 (0.68–1.83), p=0.694 | 1.50 (0.63–3.80), p=0.370 |
|  |  | 2020-2024 vs 2015-2019 | 1.40 (1.18–1.65), p=<0.001 | 1.37 (1.13–1.66), p=0.001 | 1.58 (1.07–2.36), p=0.024 | 1.09 (0.52–2.33), p=0.821 |
|  | Female | 2000-2004 vs 1995-1999 | 1.30 (0.84–2.05), p=0.257 | 1.11 (0.66–1.90), p=0.702 | 3.15 (1.03–13.69), p=0.071 | 1.60 (0.31–11.54), p=0.590 |
|  |  | 2005-2009 vs 2000-2004 | 0.47 (0.31–0.70), p=<0.001 | 0.56 (0.35–0.90), p=0.017 | 0.26 (0.08–0.67), p=0.009 | 0.42 (0.06–2.17), p=0.320 |
|  |  | 2010-2014 vs 2005-2009 | 1.55 (1.06–2.31), p=0.027 | 1.46 (0.95–2.27), p=0.089 | 1.25 (0.41–4.14), p=0.700 | 3.03 (0.73–20.39), p=0.167 |
|  |  | 2015-2019 vs 2010-2014 | 0.87 (0.65–1.18), p=0.366 | 0.84 (0.60–1.18), p=0.302 | 1.23 (0.52–3.13), p=0.640 | 1.01 (0.36–2.89), p=0.986 |
|  |  | 2020-2024 vs 2015-2019 | 1.38 (1.08–1.78), p=0.012 | 1.31 (0.98–1.75), p=0.067 | 1.68 (0.88–3.39), p=0.125 | 1.38 (0.57–3.54), p=0.477 |
|  | Male | 2000-2004 vs 1995-1999 | 0.93 (0.62–1.40), p=0.709 | 0.85 (0.50–1.46), p=0.558 | 1.37 (0.65–3.06), p=0.423 | 0.15 (0.01–0.91), p=0.080 |
|  |  | 2005-2009 vs 2000-2004 | 0.81 (0.56–1.17), p=0.258 | 1.02 (0.64–1.64), p=0.929 | 0.41 (0.18–0.86), p=0.022 | 2.46 (0.31–49.84), p=0.437 |
|  |  | 2010-2014 vs 2005-2009 | 1.21 (0.87–1.68), p=0.262 | 1.25 (0.86–1.84), p=0.245 | 1.58 (0.75–3.54), p=0.245 | 0.24 (0.01–1.85), p=0.212 |
|  |  | 2015-2019 vs 2010-2014 | 0.98 (0.75–1.29), p=0.901 | 0.91 (0.67–1.24), p=0.553 | 1.08 (0.59–2.00), p=0.800 | 5.12 (0.82–98.15), p=0.137 |
|  |  | 2020-2024 vs 2015-2019 | 1.42 (1.14–1.77), p=0.002 | 1.43 (1.11–1.85), p=0.006 | 1.54 (0.95–2.54), p=0.085 | 0.60 (0.12–2.44), p=0.480 |
| Black Caribbean | All | 2000-2004 vs 1995-1999 | 0.72 (0.58–0.89), p=0.003 | 0.82 (0.64–1.05), p=0.108 | 0.70 (0.39–1.23), p=0.219 | 0.34 (0.13–0.76), p=0.014 |
|  |  | 2005-2009 vs 2000-2004 | 0.99 (0.79–1.24), p=0.939 | 1.01 (0.79–1.29), p=0.957 | 0.93 (0.50–1.71), p=0.806 | 1.13 (0.40–3.25), p=0.812 |
|  |  | 2010-2014 vs 2005-2009 | 1.05 (0.85–1.30), p=0.644 | 1.11 (0.88–1.39), p=0.397 | 0.76 (0.40–1.44), p=0.403 | 1.17 (0.46–3.08), p=0.741 |
|  |  | 2015-2019 vs 2010-2014 | 1.08 (0.89–1.32), p=0.435 | 1.08 (0.87–1.33), p=0.483 | 1.35 (0.73–2.56), p=0.337 | 0.58 (0.21–1.52), p=0.275 |
|  |  | 2020-2024 vs 2015-2019 | 1.10 (0.92–1.32), p=0.289 | 0.98 (0.80–1.20), p=0.849 | 1.75 (1.09–2.89), p=0.024 | 2.06 (0.88–5.36), p=0.112 |
|  | Female | 2000-2004 vs 1995-1999 | 0.74 (0.53–1.01), p=0.056 | 0.89 (0.62–1.28), p=0.528 | 0.35 (0.10–1.02), p=0.071 | 0.50 (0.17–1.32), p=0.176 |
|  |  | 2005-2009 vs 2000-2004 | 1.02 (0.74–1.40), p=0.920 | 0.93 (0.65–1.33), p=0.674 | 2.94 (1.03–10.48), p=0.061 | 0.84 (0.24–2.80), p=0.772 |
|  |  | 2010-2014 vs 2005-2009 | 1.01 (0.75–1.36), p=0.971 | 1.12 (0.80–1.57), p=0.502 | 0.43 (0.15–1.08), p=0.084 | 1.24 (0.39–4.20), p=0.714 |
|  |  | 2015-2019 vs 2010-2014 | 1.09 (0.82–1.44), p=0.549 | 1.04 (0.77–1.42), p=0.785 | 2.45 (1.00–6.85), p=0.063 | 0.47 (0.12–1.57), p=0.232 |
|  |  | 2020-2024 vs 2015-2019 | 1.11 (0.86–1.43), p=0.432 | 1.02 (0.77–1.36), p=0.891 | 1.09 (0.56–2.16), p=0.792 | 2.87 (1.01–10.22), p=0.065 |
|  | Male | 2000-2004 vs 1995-1999 | 0.72 (0.54–0.96), p=0.028 | 0.77 (0.55–1.08), p=0.127 | 0.94 (0.47–1.85), p=0.846 | 0.12 (0.01–0.64), p=0.044 |
|  |  | 2005-2009 vs 2000-2004 | 0.98 (0.72–1.33), p=0.878 | 1.10 (0.78–1.55), p=0.605 | 0.45 (0.18–1.01), p=0.062 | 3.03 (0.38–61.81), p=0.342 |
|  |  | 2010-2014 vs 2005-2009 | 1.11 (0.82–1.49), p=0.496 | 1.10 (0.80–1.52), p=0.554 | 1.34 (0.54–3.47), p=0.533 | 1.05 (0.19–5.71), p=0.953 |
|  |  | 2015-2019 vs 2010-2014 | 1.09 (0.83–1.43), p=0.553 | 1.13 (0.84–1.52), p=0.427 | 0.77 (0.31–1.87), p=0.564 | 0.84 (0.15–4.57), p=0.832 |
|  |  | 2020-2024 vs 2015-2019 | 1.11 (0.86–1.43), p=0.430 | 0.95 (0.72–1.27), p=0.742 | 2.96 (1.45–6.66), p=0.005 | 0.91 (0.17–4.96), p=0.909 |
| Other | All | 2000-2004 vs 1995-1999 | 0.85 (0.63–1.16), p=0.305 | 0.80 (0.55–1.17), p=0.252 | 0.78 (0.41–1.48), p=0.439 | 1.31 (0.49–3.85), p=0.602 |
|  |  | 2005-2009 vs 2000-2004 | 0.65 (0.48–0.87), p=0.004 | 0.72 (0.50–1.04), p=0.079 | 0.59 (0.30–1.14), p=0.117 | 0.34 (0.11–0.96), p=0.049 |
|  |  | 2010-2014 vs 2005-2009 | 0.90 (0.67–1.19), p=0.449 | 1.01 (0.73–1.41), p=0.958 | 0.74 (0.37–1.45), p=0.373 | 0.59 (0.15–2.24), p=0.437 |
|  |  | 2015-2019 vs 2010-2014 | 0.83 (0.64–1.09), p=0.176 | 0.74 (0.54–1.00), p=0.052 | 0.95 (0.50–1.83), p=0.888 | 2.37 (0.81–8.56), p=0.140 |
|  |  | 2020-2024 vs 2015-2019 | 1.52 (1.21–1.92), p=<0.001 | 1.56 (1.19–2.06), p=0.001 | 1.65 (0.99–2.85), p=0.062 | 0.96 (0.42–2.21), p=0.921 |
|  | Female | 2000-2004 vs 1995-1999 | 0.77 (0.48–1.24), p=0.276 | 0.81 (0.44–1.50), p=0.489 | 0.60 (0.22–1.62), p=0.314 | 0.92 (0.28–3.18), p=0.884 |
|  |  | 2005-2009 vs 2000-2004 | 0.82 (0.52–1.29), p=0.385 | 0.88 (0.50–1.57), p=0.667 | 0.94 (0.36–2.59), p=0.898 | 0.33 (0.07–1.27), p=0.121 |
|  |  | 2010-2014 vs 2005-2009 | 0.84 (0.56–1.28), p=0.412 | 1.03 (0.64–1.68), p=0.907 | 0.50 (0.18–1.31), p=0.164 | 0.49 (0.06–2.95), p=0.434 |
|  |  | 2015-2019 vs 2010-2014 | 0.85 (0.58–1.25), p=0.398 | 0.76 (0.49–1.18), p=0.224 | 0.86 (0.31–2.47), p=0.777 | 2.59 (0.60–17.67), p=0.245 |
|  |  | 2020-2024 vs 2015-2019 | 1.37 (0.98–1.93), p=0.065 | 1.34 (0.91–1.99), p=0.147 | 1.94 (0.87–4.73), p=0.119 | 0.87 (0.27–2.80), p=0.816 |
|  | Male | 2000-2004 vs 1995-1999 | 0.92 (0.62–1.37), p=0.676 | 0.80 (0.50–1.29), p=0.368 | 0.95 (0.41–2.25), p=0.906 | 3.12 (0.46–61.03), p=0.310 |
|  |  | 2005-2009 vs 2000-2004 | 0.55 (0.37–0.82), p=0.003 | 0.63 (0.39–1.02), p=0.061 | 0.41 (0.15–1.01), p=0.059 | 0.33 (0.05–1.71), p=0.205 |
|  |  | 2010-2014 vs 2005-2009 | 0.95 (0.64–1.42), p=0.805 | 0.99 (0.63–1.57), p=0.978 | 1.09 (0.42–3.01), p=0.860 | 0.76 (0.09–6.32), p=0.781 |
|  |  | 2015-2019 vs 2010-2014 | 0.81 (0.56–1.17), p=0.260 | 0.71 (0.46–1.09), p=0.114 | 1.00 (0.44–2.36), p=0.991 | 2.12 (0.45–14.82), p=0.371 |
|  |  | 2020-2024 vs 2015-2019 | 1.66 (1.21–2.30), p=0.002 | 1.80 (1.23–2.66), p=0.003 | 1.46 (0.75–2.99), p=0.276 | 1.05 (0.32–3.64), p=0.937 |

**Supplementary Table 5.** Age-, sex-adjusted incidence rate ratios with 95% confidence intervals and p-values for first-ever stroke by ethnic group, stroke subtype, and time period in the total population, and age-adjusted IRRs stratified by sex (female and male populations), South London Stroke Register, 1995–2024

| **Period** | **Ethnicity** | **All Strokes** | **Ischaemic Strokes** | **PICH** | **SAH** |
| --- | --- | --- | --- | --- | --- |
| **Total Population** | | | | | |
| 1995-1999 | Black African | 1.57 (1.23–2.00), p=<0.001 | 1.60 (1.19–2.15), p=0.002 | 2.18 (1.24–3.82), p=0.007 | 1.32 (0.62–2.82), p=0.469 |
|  | Black Caribbean | 1.52 (1.31–1.77), p=<0.001 | 1.46 (1.22–1.75), p=<0.001 | 1.83 (1.23–2.74), p=0.003 | 2.95 (1.80–4.85), p=<0.001 |
|  | Other | 1.15 (0.92–1.44), p=0.224 | 1.11 (0.84–1.46), p=0.466 | 2.16 (1.35–3.46), p=0.001 | 1.14 (0.52–2.52), p=0.747 |
| 2000-2004 | Black African | 1.76 (1.44–2.15), p=<0.001 | 1.63 (1.27–2.10), p=<0.001 | 3.67 (2.46–5.47), p=<0.001 | 0.60 (0.26–1.41), p=0.244 |
|  | Black Caribbean | 1.21 (1.02–1.43), p=0.031 | 1.25 (1.03–1.52), p=0.021 | 1.43 (0.91–2.27), p=0.124 | 0.91 (0.44–1.91), p=0.805 |
|  | Other | 1.06 (0.85–1.31), p=0.627 | 0.94 (0.71–1.23), p=0.634 | 1.76 (1.09–2.84), p=0.021 | 1.23 (0.65–2.33), p=0.529 |
| 2005-2009 | Black African | 1.31 (1.05–1.62), p=0.016 | 1.24 (0.97–1.58), p=0.087 | 2.04 (1.18–3.53), p=0.011 | 1.46 (0.60–3.54), p=0.404 |
|  | Black Caribbean | 1.41 (1.19–1.67), p=<0.001 | 1.35 (1.12–1.63), p=0.002 | 1.97 (1.23–3.17), p=0.005 | 2.43 (1.16–5.09), p=0.019 |
|  | Other | 0.84 (0.67–1.05), p=0.120 | 0.72 (0.56–0.94), p=0.015 | 1.68 (1.00–2.81), p=0.049 | 1.22 (0.51–2.93), p=0.649 |
| 2010-2014 | Black African | 1.92 (1.62–2.28), p=<0.001 | 1.81 (1.50–2.19), p=<0.001 | 2.63 (1.69–4.11), p=<0.001 | 1.90 (0.89–4.08), p=0.099 |
|  | Black Caribbean | 1.74 (1.48–2.05), p=<0.001 | 1.71 (1.44–2.05), p=<0.001 | 1.59 (0.94–2.67), p=0.082 | 3.22 (1.59–6.54), p=0.001 |
|  | Other | 0.86 (0.70–1.05), p=0.140 | 0.82 (0.65–1.03), p=0.086 | 1.25 (0.74–2.09), p=0.404 | 0.78 (0.30–2.02), p=0.604 |
| 2015-2019 | Black African | 1.84 (1.57–2.15), p=<0.001 | 1.81 (1.52–2.16), p=<0.001 | 1.98 (1.36–2.89), p=<0.001 | 1.88 (1.00–3.54), p=0.051 |
|  | Black Caribbean | 1.92 (1.65–2.24), p=<0.001 | 1.98 (1.67–2.34), p=<0.001 | 1.63 (1.05–2.52), p=0.028 | 1.67 (0.77–3.64), p=0.198 |
|  | Other | 0.74 (0.60–0.90), p=0.003 | 0.67 (0.53–0.84), p=<0.001 | 0.90 (0.57–1.44), p=0.665 | 1.34 (0.69–2.60), p=0.383 |
| 2020-2024 | Black African | 2.31 (2.03–2.62), p=<0.001 | 2.22 (1.93–2.57), p=<0.001 | 3.04 (2.24–4.12), p=<0.001 | 1.65 (0.92–2.95), p=0.092 |
|  | Black Caribbean | 2.00 (1.73–2.31), p=<0.001 | 1.86 (1.58–2.19), p=<0.001 | 2.59 (1.83–3.66), p=<0.001 | 2.61 (1.47–4.63), p=0.001 |
|  | Other | 1.04 (0.89–1.22), p=0.625 | 0.97 (0.81–1.16), p=0.744 | 1.45 (1.01–2.08), p=0.044 | 1.09 (0.59–2.04), p=0.782 |
| **Female Population** | | | | | |
| 1995-1999 | Black African | 1.61 (1.10–2.35), p=0.014 | 2.04 (1.33–3.13), p=0.001 | 1.48 (0.52–4.19), p=0.461 | 0.81 (0.23–2.81), p=0.739 |
|  | Black Caribbean | 1.58 (1.26–1.98), p=<0.001 | 1.53 (1.16–2.01), p=0.002 | 1.91 (1.03–3.53), p=0.040 | 2.91 (1.52–5.58), p=0.001 |
|  | Other | 1.18 (0.84–1.67), p=0.336 | 1.05 (0.68–1.64), p=0.816 | 2.82 (1.45–5.47), p=0.002 | 1.79 (0.75–4.29), p=0.193 |
| 2000-2004 | Black African | 2.05 (1.54–2.72), p=<0.001 | 2.09 (1.49–2.94), p=<0.001 | 4.15 (2.26–7.61), p=<0.001 | 0.88 (0.34–2.32), p=0.803 |
|  | Black Caribbean | 1.22 (0.95–1.57), p=0.116 | 1.33 (1.01–1.75), p=0.043 | 0.73 (0.28–1.87), p=0.511 | 1.44 (0.64–3.26), p=0.378 |
|  | Other | 0.96 (0.68–1.36), p=0.810 | 0.83 (0.54–1.29), p=0.415 | 1.79 (0.84–3.79), p=0.130 | 1.53 (0.67–3.46), p=0.312 |
| 2005-2009 | Black African | 1.20 (0.85–1.68), p=0.293 | 1.25 (0.86–1.82), p=0.242 | 1.71 (0.72–4.07), p=0.222 | 0.91 (0.26–3.14), p=0.875 |
|  | Black Caribbean | 1.48 (1.16–1.88), p=0.001 | 1.38 (1.05–1.82), p=0.019 | 2.32 (1.27–4.25), p=0.006 | 2.18 (0.88–5.38), p=0.090 |
|  | Other | 0.98 (0.71–1.34), p=0.882 | 0.82 (0.56–1.20), p=0.309 | 2.32 (1.20–4.50), p=0.013 | 1.17 (0.40–3.39), p=0.773 |
| 2010-2014 | Black African | 2.01 (1.56–2.59), p=<0.001 | 2.02 (1.52–2.68), p=<0.001 | 1.49 (0.72–3.10), p=0.285 | 2.65 (1.12–6.28), p=0.027 |
|  | Black Caribbean | 1.73 (1.37–2.19), p=<0.001 | 1.78 (1.38–2.29), p=<0.001 | 1.02 (0.45–2.28), p=0.969 | 3.52 (1.50–8.26), p=0.004 |
|  | Other | 0.94 (0.69–1.27), p=0.669 | 0.96 (0.69–1.34), p=0.820 | 1.03 (0.48–2.19), p=0.948 | 0.72 (0.21–2.53), p=0.610 |
| 2015-2019 | Black African | 1.78 (1.41–2.25), p=<0.001 | 1.84 (1.42–2.40), p=<0.001 | 1.48 (0.81–2.71), p=0.204 | 1.89 (0.86–4.17), p=0.113 |
|  | Black Caribbean | 1.99 (1.59–2.48), p=<0.001 | 2.03 (1.59–2.60), p=<0.001 | 2.02 (1.15–3.54), p=0.014 | 1.47 (0.55–3.92), p=0.447 |
|  | Other | 0.83 (0.62–1.11), p=0.202 | 0.81 (0.58–1.13), p=0.209 | 0.79 (0.38–1.61), p=0.510 | 1.25 (0.53–2.93), p=0.612 |
| 2020-2024 | Black African | 2.07 (1.71–2.51), p=<0.001 | 1.99 (1.60–2.47), p=<0.001 | 2.36 (1.47–3.78), p=<0.001 | 2.49 (1.23–5.04), p=0.011 |
|  | Black Caribbean | 1.92 (1.56–2.35), p=<0.001 | 1.79 (1.42–2.26), p=<0.001 | 1.96 (1.16–3.31), p=0.012 | 4.00 (2.00–7.99), p=<0.001 |
|  | Other | 0.98 (0.77–1.24), p=0.862 | 0.92 (0.70–1.20), p=0.524 | 1.39 (0.82–2.36), p=0.221 | 1.09 (0.46–2.59), p=0.837 |
| **Male Population** | | | | | |
| 1995-1999 | Black African | 1.57 (1.14–2.16), p=0.006 | 1.38 (0.92–2.06), p=0.120 | 2.74 (1.44–5.20), p=0.002 | 2.10 (0.86–5.11), p=0.103 |
|  | Black Caribbean | 1.52 (1.23–1.86), p=<0.001 | 1.45 (1.14–1.85), p=0.002 | 1.86 (1.11–3.11), p=0.018 | 3.20 (1.57–6.53), p=0.001 |
|  | Other | 1.14 (0.85–1.54), p=0.387 | 1.16 (0.82–1.64), p=0.395 | 1.86 (0.99–3.49), p=0.055 | 0.56 (0.12–2.63), p=0.461 |
| 2000-2004 | Black African | 1.56 (1.17–2.07), p=0.002 | 1.32 (0.91–1.90), p=0.141 | 3.44 (2.06–5.76), p=<0.001 | 0.37 (0.08–1.70), p=0.200 |
|  | Black Caribbean | 1.23 (0.97–1.56), p=0.084 | 1.24 (0.94–1.62), p=0.124 | 2.00 (1.18–3.37), p=0.009 | 0.39 (0.08–1.80), p=0.227 |
|  | Other | 1.15 (0.87–1.53), p=0.327 | 1.04 (0.73–1.46), p=0.839 | 1.83 (1.01–3.31), p=0.048 | 1.03 (0.41–2.60), p=0.953 |
| 2005-2009 | Black African | 1.44 (1.09–1.91), p=0.010 | 1.28 (0.93–1.76), p=0.129 | 2.53 (1.29–4.97), p=0.007 | 2.84 (0.96–8.46), p=0.060 |
|  | Black Caribbean | 1.41 (1.11–1.79), p=0.005 | 1.38 (1.06–1.78), p=0.015 | 1.84 (0.88–3.82), p=0.103 | 2.88 (0.96–8.66), p=0.060 |
|  | Other | 0.77 (0.56–1.05), p=0.097 | 0.69 (0.48–0.98), p=0.038 | 1.34 (0.63–2.86), p=0.451 | 1.46 (0.43–4.96), p=0.541 |
| 2010-2014 | Black African | 1.90 (1.51–2.40), p=<0.001 | 1.72 (1.33–2.23), p=<0.001 | 3.97 (2.28–6.90), p=<0.001 | 0.92 (0.20–4.16), p=0.915 |
|  | Black Caribbean | 1.84 (1.46–2.31), p=<0.001 | 1.76 (1.38–2.25), p=<0.001 | 2.42 (1.25–4.69), p=0.009 | 3.12 (1.05–9.26), p=0.041 |
|  | Other | 0.83 (0.63–1.10), p=0.198 | 0.75 (0.55–1.03), p=0.079 | 1.55 (0.79–3.03), p=0.204 | 1.08 (0.32–3.59), p=0.906 |
| 2015-2019 | Black African | 1.90 (1.55–2.34), p=<0.001 | 1.80 (1.42–2.27), p=<0.001 | 2.50 (1.56–4.00), p=<0.001 | 2.01 (0.79–5.12), p=0.144 |
|  | Black Caribbean | 1.89 (1.53–2.34), p=<0.001 | 1.96 (1.56–2.47), p=<0.001 | 1.26 (0.64–2.50), p=0.505 | 2.33 (0.78–6.99), p=0.130 |
|  | Other | 0.67 (0.51–0.89), p=0.005 | 0.57 (0.41–0.79), p=<0.001 | 1.04 (0.58–1.87), p=0.893 | 1.59 (0.63–4.01), p=0.321 |
| 2020-2024 | Black African | 2.52 (2.13–2.98), p=<0.001 | 2.43 (2.00–2.94), p=<0.001 | 3.65 (2.47–5.40), p=<0.001 | 0.82 (0.28–2.38), p=0.716 |
|  | Black Caribbean | 2.10 (1.72–2.57), p=<0.001 | 1.96 (1.56–2.47), p=<0.001 | 3.29 (2.10–5.16), p=<0.001 | 1.22 (0.42–3.57), p=0.718 |
|  | Other | 1.10 (0.89–1.36), p=0.387 | 1.02 (0.80–1.31), p=0.851 | 1.51 (0.93–2.45), p=0.093 | 1.21 (0.53–2.75), p=0.650 |

**Supplementary Table 6.** Age-, sex-, and IMD-adjusted incidence rate ratios with 95% confidence intervals and p-values for first-ever stroke by ethnic group, stroke subtype, and time period in the total population, and age- and IMD-adjusted IRRs stratified by sex (female and male populations), South London Stroke Register, 1995–2024 (Reference= White on the same period)

| **Period** | **Ethnicity** | **All Strokes** | **Ischaemic Strokes** | **PICH** | **SAH** |
| --- | --- | --- | --- | --- | --- |
| **Total Population** | | | | | |
| 1995-1999 | Black African | 1.67 (1.31–2.14), p=<0.001 | 1.70 (1.27–2.29), p=<0.001 | 2.28 (1.30–4.00), p=0.004 | 1.47 (0.69–3.13), p=0.316 |
|  | Black Caribbean | 1.65 (1.41–1.93), p=<0.001 | 1.59 (1.32–1.92), p=<0.001 | 1.98 (1.32–2.96), p=<0.001 | 3.33 (2.01–5.51), p=<0.001 |
|  | Other | 1.10 (0.87–1.38), p=0.432 | 1.06 (0.80–1.40), p=0.680 | 2.02 (1.25–3.28), p=0.004 | 1.10 (0.50–2.42), p=0.814 |
| 2000-2004 | Black African | 1.85 (1.51–2.27), p=<0.001 | 1.73 (1.34–2.23), p=<0.001 | 3.72 (2.48–5.58), p=<0.001 | 0.65 (0.28–1.53), p=0.323 |
|  | Black Caribbean | 1.26 (1.06–1.51), p=0.010 | 1.35 (1.11–1.64), p=0.003 | 1.32 (0.81–2.16), p=0.261 | 1.00 (0.48–2.09), p=0.996 |
|  | Other | 1.08 (0.87–1.35), p=0.481 | 0.96 (0.73–1.26), p=0.774 | 1.79 (1.11–2.89), p=0.017 | 1.27 (0.67–2.41), p=0.456 |
| 2005-2009 | Black African | 1.27 (1.02–1.58), p=0.032 | 1.21 (0.94–1.55), p=0.134 | 1.96 (1.13–3.39), p=0.017 | 1.43 (0.59–3.47), p=0.426 |
|  | Black Caribbean | 1.31 (1.10–1.55), p=0.002 | 1.25 (1.03–1.51), p=0.021 | 1.83 (1.14–2.94), p=0.013 | 2.34 (1.12–4.93), p=0.025 |
|  | Other | 0.90 (0.72–1.12), p=0.335 | 0.78 (0.60–1.01), p=0.059 | 1.76 (1.05–2.95), p=0.031 | 1.26 (0.53–2.99), p=0.599 |
| 2010-2014 | Black African | 1.71 (1.44–2.03), p=<0.001 | 1.62 (1.33–1.96), p=<0.001 | 2.31 (1.48–3.62), p=<0.001 | 1.60 (0.75–3.42), p=0.228 |
|  | Black Caribbean | 1.54 (1.31–1.82), p=<0.001 | 1.52 (1.27–1.82), p=<0.001 | 1.38 (0.82–2.33), p=0.222 | 2.68 (1.32–5.44), p=0.006 |
|  | Other | 0.81 (0.65–0.99), p=0.042 | 0.77 (0.61–0.97), p=0.026 | 1.18 (0.70–1.98), p=0.531 | 0.72 (0.28–1.87), p=0.502 |
| 2015-2019 | Black African | 1.60 (1.37–1.87), p=<0.001 | 1.57 (1.32–1.87), p=<0.001 | 1.77 (1.21–2.58), p=0.003 | 1.62 (0.86–3.05), p=0.137 |
|  | Black Caribbean | 1.78 (1.52–2.08), p=<0.001 | 1.82 (1.54–2.16), p=<0.001 | 1.53 (0.99–2.37), p=0.057 | 1.50 (0.69–3.27), p=0.303 |
|  | Other | 0.69 (0.56–0.84), p=<0.001 | 0.62 (0.49–0.79), p=<0.001 | 0.85 (0.54–1.35), p=0.498 | 1.24 (0.64–2.39), p=0.527 |
| 2020-2024 | Black African | 2.03 (1.78–2.32), p=<0.001 | 1.92 (1.66–2.23), p=<0.001 | 3.00 (2.18–4.13), p=<0.001 | 1.27 (0.68–2.35), p=0.456 |
|  | Black Caribbean | 1.79 (1.54–2.07), p=<0.001 | 1.68 (1.43–1.99), p=<0.001 | 2.44 (1.69–3.53), p=<0.001 | 1.85 (0.96–3.56), p=0.064 |
|  | Other | 0.96 (0.82–1.13), p=0.654 | 0.89 (0.74–1.07), p=0.218 | 1.40 (0.96–2.06), p=0.083 | 1.05 (0.56–1.97), p=0.873 |
| **Female Population** | | | | | |
| 1995-1999 | Black African | 1.68 (1.15–2.46), p=0.007 | 2.11 (1.37–3.25), p=<0.001 | 1.48 (0.52–4.18), p=0.460 | 0.91 (0.26–3.14), p=0.880 |
|  | Black Caribbean | 1.73 (1.38–2.19), p=<0.001 | 1.69 (1.28–2.23), p=<0.001 | 1.99 (1.07–3.69), p=0.029 | 3.26 (1.69–6.28), p=<0.001 |
|  | Other | 1.11 (0.78–1.56), p=0.567 | 0.97 (0.62–1.51), p=0.893 | 2.73 (1.41–5.31), p=0.003 | 1.81 (0.76–4.31), p=0.183 |
| 2000-2004 | Black African | 2.12 (1.59–2.83), p=<0.001 | 2.14 (1.52–3.02), p=<0.001 | 4.26 (2.33–7.79), p=<0.001 | 1.00 (0.38–2.62), p=0.999 |
|  | Black Caribbean | 1.28 (0.99–1.65), p=0.055 | 1.41 (1.07–1.87), p=0.015 | 0.76 (0.30–1.94), p=0.566 | 1.65 (0.72–3.74), p=0.234 |
|  | Other | 0.95 (0.67–1.34), p=0.765 | 0.82 (0.53–1.27), p=0.381 | 1.75 (0.83–3.69), p=0.142 | 1.58 (0.70–3.57), p=0.269 |
| 2005-2009 | Black African | 1.19 (0.85–1.67), p=0.321 | 1.24 (0.85–1.80), p=0.262 | 1.68 (0.71–3.99), p=0.236 | 0.94 (0.27–3.21), p=0.917 |
|  | Black Caribbean | 1.43 (1.12–1.82), p=0.004 | 1.33 (1.01–1.75), p=0.040 | 2.24 (1.22–4.08), p=0.009 | 2.26 (0.92–5.57), p=0.076 |
|  | Other | 1.05 (0.76–1.45), p=0.753 | 0.89 (0.61–1.31), p=0.554 | 2.49 (1.29–4.81), p=0.007 | 1.18 (0.42–3.36), p=0.752 |
| 2010-2014 | Black African | 1.77 (1.37–2.29), p=<0.001 | 1.78 (1.33–2.37), p=<0.001 | 1.33 (0.64–2.76), p=0.445 | 2.29 (0.97–5.39), p=0.058 |
|  | Black Caribbean | 1.56 (1.23–1.98), p=<0.001 | 1.60 (1.24–2.07), p=<0.001 | 0.91 (0.41–2.05), p=0.829 | 3.03 (1.30–7.08), p=0.010 |
|  | Other | 0.89 (0.65–1.20), p=0.438 | 0.91 (0.65–1.27), p=0.577 | 0.98 (0.46–2.09), p=0.968 | 0.68 (0.20–2.35), p=0.547 |
| 2015-2019 | Black African | 1.56 (1.24–1.98), p=<0.001 | 1.63 (1.25–2.12), p=<0.001 | 1.30 (0.71–2.38), p=0.398 | 1.60 (0.73–3.51), p=0.242 |
|  | Black Caribbean | 1.84 (1.48–2.30), p=<0.001 | 1.89 (1.48–2.42), p=<0.001 | 1.89 (1.08–3.31), p=0.026 | 1.29 (0.49–3.42), p=0.609 |
|  | Other | 0.77 (0.58–1.04), p=0.085 | 0.75 (0.54–1.05), p=0.099 | 0.74 (0.36–1.51), p=0.411 | 1.15 (0.49–2.67), p=0.751 |
| 2020-2024 | Black African | 1.83 (1.50–2.23), p=<0.001 | 1.73 (1.38–2.17), p=<0.001 | 2.22 (1.35–3.66), p=0.002 | 2.07 (0.99–4.32), p=0.053 |
|  | Black Caribbean | 1.71 (1.38–2.12), p=<0.001 | 1.66 (1.31–2.11), p=<0.001 | 1.76 (1.00–3.09), p=0.052 | 2.64 (1.19–5.89), p=0.017 |
|  | Other | 0.94 (0.74–1.19), p=0.611 | 0.86 (0.65–1.14), p=0.290 | 1.42 (0.82–2.47), p=0.207 | 1.08 (0.46–2.57), p=0.855 |
| **Male Population** | | | | | |
| 1995-1999 | Black African | 1.68 (1.22–2.32), p=0.001 | 1.48 (0.99–2.22), p=0.055 | 2.93 (1.55–5.56), p=<0.001 | 2.34 (0.96–5.69), p=0.061 |
|  | Black Caribbean | 1.62 (1.31–2.00), p=<0.001 | 1.55 (1.20–1.99), p=<0.001 | 2.05 (1.22–3.44), p=0.007 | 3.67 (1.78–7.57), p=<0.001 |
|  | Other | 1.09 (0.80–1.49), p=0.571 | 1.13 (0.79–1.62), p=0.491 | 1.67 (0.87–3.23), p=0.125 | 0.52 (0.11–2.40), p=0.401 |
| 2000-2004 | Black African | 1.66 (1.24–2.21), p=<0.001 | 1.44 (1.00–2.08), p=0.051 | 3.46 (2.05–5.85), p=<0.001 | 0.38 (0.08–1.74), p=0.212 |
|  | Black Caribbean | 1.28 (1.00–1.64), p=0.048 | 1.34 (1.01–1.76), p=0.040 | 1.83 (1.04–3.23), p=0.037 | 0.41 (0.09–1.88), p=0.249 |
|  | Other | 1.21 (0.91–1.60), p=0.187 | 1.09 (0.77–1.55), p=0.610 | 1.89 (1.05–3.43), p=0.035 | 1.07 (0.43–2.67), p=0.893 |
| 2005-2009 | Black African | 1.37 (1.03–1.82), p=0.028 | 1.22 (0.89–1.69), p=0.220 | 2.34 (1.19–4.59), p=0.014 | 2.50 (0.84–7.43), p=0.098 |
|  | Black Caribbean | 1.25 (0.98–1.59), p=0.067 | 1.23 (0.95–1.59), p=0.119 | 1.62 (0.78–3.37), p=0.194 | 2.43 (0.81–7.24), p=0.112 |
|  | Other | 0.81 (0.60–1.11), p=0.187 | 0.73 (0.51–1.04), p=0.080 | 1.36 (0.64–2.90), p=0.419 | 1.53 (0.46–5.02), p=0.487 |
| 2010-2014 | Black African | 1.70 (1.34–2.14), p=<0.001 | 1.55 (1.19–2.00), p=<0.001 | 3.43 (1.97–5.98), p=<0.001 | 0.74 (0.17–3.29), p=0.696 |
|  | Black Caribbean | 1.60 (1.27–2.02), p=<0.001 | 1.54 (1.20–1.98), p=<0.001 | 2.05 (1.06–3.98), p=0.034 | 2.46 (0.84–7.23), p=0.102 |
|  | Other | 0.78 (0.59–1.03), p=0.083 | 0.71 (0.51–0.97), p=0.031 | 1.45 (0.74–2.83), p=0.282 | 0.97 (0.30–3.17), p=0.963 |
| 2015-2019 | Black African | 1.64 (1.33–2.02), p=<0.001 | 1.54 (1.21–1.95), p=<0.001 | 2.26 (1.40–3.62), p=<0.001 | 1.78 (0.70–4.53), p=0.223 |
|  | Black Caribbean | 1.74 (1.40–2.16), p=<0.001 | 1.80 (1.43–2.27), p=<0.001 | 1.19 (0.60–2.35), p=0.620 | 2.18 (0.74–6.46), p=0.158 |
|  | Other | 0.63 (0.48–0.83), p=0.001 | 0.54 (0.39–0.75), p=<0.001 | 0.98 (0.55–1.76), p=0.955 | 1.47 (0.59–3.67), p=0.408 |
| 2020-2024 | Black African | 2.21 (1.85–2.62), p=<0.001 | 2.09 (1.72–2.53), p=<0.001 | 3.65 (2.44–5.48), p=<0.001 | 0.50 (0.15–1.70), p=0.269 |
|  | Black Caribbean | 1.88 (1.53–2.31), p=<0.001 | 1.73 (1.37–2.19), p=<0.001 | 3.17 (1.98–5.07), p=<0.001 | 1.19 (0.41–3.44), p=0.751 |
|  | Other | 0.99 (0.79–1.23), p=0.909 | 0.92 (0.72–1.18), p=0.514 | 1.38 (0.83–2.31), p=0.217 | 1.13 (0.50–2.56), p=0.767 |

**Supplementary Table 7.** Interaction analysis yielding Age-, sex-, and SES-adjusted IRRs for first-ever stroke by ethnic group, sex, and IMD quintile, stratified by stroke subtype, South London Stroke Register, 1995–2024.

|  |  |  | **All-strokes** | | **Ischaemic Strokes** | | **PICH Strokes** | | **SAH strokes** | |
| --- | --- | --- | --- | --- | --- | --- | --- | --- | --- | --- |
| **Ethnicity** | **Sex** | **IMD** | **IRR (95%CI)** | **p value** | **IRR (95%CI)** | **p value** | **IRR (95%CI)** | **p value** | **IRR (95%CI)** | **p value** |
| White | All | 1 | 1.55 (1.42–1.71) | <0.001 | 1.52 (1.37–1.68) | <0.001 | 1.29 (1.01–1.66) | 0.045 | 1.97 (1.27–3.05) | 0.003 |
|  |  | 2 | 0.91 (0.82–1.00) | 0.043 | 0.91 (0.82–1.02) | 0.105 | 0.75 (0.58–0.98) | 0.035 | 0.89 (0.56–1.42) | 0.637 |
|  |  | ≥3 | 1.00 (1.00–1.00) |  | 1.00 (1.00–1.00) | Reference | 1.00 (1.00–1.00) |  | 1.00 (1.00–1.00) |  |
|  | Female | 1 | 1.45 (1.27–1.66) | <0.001 | 1.39 (1.20–1.62) | <0.001 | 1.37 (0.93–2.01) | 0.111 | 2.08 (1.09–3.97) | 0.026 |
|  |  | 2 | 0.85 (0.74–0.98) | 0.022 | 0.83 (0.71–0.98) | 0.024 | 0.77 (0.52–1.16) | 0.212 | 1.08 (0.55–2.12) | 0.817 |
|  |  | ≥3 | 1.00 (1.00–1.00) |  | 1.00 (1.00–1.00) | Reference | 1.00 (1.00–1.00) |  | 1.00 (1.00–1.00) |  |
|  | Male | 1 | 1.64 (1.44–1.86) | <0.001 | 1.63 (1.41–1.88) | <0.001 | 1.22 (0.88–1.70) | 0.239 | 1.89 (1.03–3.46) | 0.039 |
|  |  | 2 | 0.95 (0.83–1.08) | 0.440 | 0.98 (0.85–1.14) | 0.790 | 0.73 (0.52–1.04) | 0.079 | 0.74 (0.38–1.41) | 0.356 |
|  |  | ≥3 | 1.00 (1.00–1.00) |  | 1.00 (1.00–1.00) | Reference | 1.00 (1.00–1.00) |  | 1.00 (1.00–1.00) |  |
| Black African | All | 1 | 2.76 (2.45–3.12) | <0.001 | 2.73 (2.38–3.13) | <0.001 | 3.28 (2.43–4.42) | <0.001 | 2.06 (1.18–3.57) | 0.010 |
|  |  | 2 | 1.63 (1.43–1.85) | <0.001 | 1.58 (1.37–1.83) | <0.001 | 2.05 (1.50–2.80) | <0.001 | 1.36 (0.75–2.46) | 0.312 |
|  |  | ≥3 | 1.18 (0.92–1.51) | 0.193 | 1.07 (0.80–1.43) | 0.659 | 1.74 (1.02–2.99) | 0.043 | 1.22 (0.42–3.52) | 0.713 |
|  | Female | 1 | 2.40 (2.00–2.87) | <0.001 | 2.40 (1.96–2.94) | <0.001 | 2.80 (1.73–4.52) | <0.001 | 2.48 (1.16–5.30) | 0.019 |
|  |  | 2 | 1.54 (1.28–1.86) | <0.001 | 1.56 (1.26–1.93) | <0.001 | 1.58 (0.94–2.66) | 0.085 | 1.96 (0.90–4.27) | 0.092 |
|  |  | ≥3 | 1.08 (0.74–1.56) | 0.695 | 1.04 (0.67–1.60) | 0.864 | 1.32 (0.51–3.38) | 0.569 | 1.17 (0.26–5.28) | 0.839 |
|  | Male | 1 | 3.10 (2.64–3.64) | <0.001 | 3.02 (2.52–3.63) | <0.001 | 3.64 (2.48–5.33) | <0.001 | 1.65 (0.72–3.78) | 0.233 |
|  |  | 2 | 1.73 (1.45–2.06) | <0.001 | 1.62 (1.33–1.98) | <0.001 | 2.42 (1.63–3.60) | <0.001 | 0.74 (0.26–2.08) | 0.567 |
|  |  | ≥3 | 1.27 (0.92–1.76) | 0.151 | 1.10 (0.74–1.63) | 0.631 | 2.07 (1.07–4.01) | 0.030 | 1.30 (0.29–5.77) | 0.730 |
| Black Caribbean | All | 1 | 2.20 (1.96–2.47) | <0.001 | 2.14 (1.88–2.44) | <0.001 | 2.28 (1.68–3.10) | <0.001 | 2.99 (1.77–5.07) | <0.001 |
|  |  | 2 | 1.53 (1.35–1.73) | <0.001 | 1.53 (1.33–1.76) | <0.001 | 1.40 (1.00–1.98) | 0.052 | 2.49 (1.43–4.32) | 0.001 |
|  |  | ≥3 | 1.44 (1.13–1.84) | 0.003 | 1.55 (1.20–2.01) | <0.001 | 0.98 (0.45–2.12) | 0.958 | 1.51 (0.45–5.00) | 0.504 |
|  | Female | 1 | 1.97 (1.66–2.33) | <0.001 | 1.92 (1.59–2.33) | <0.001 | 2.11 (1.31–3.41) | 0.002 | 3.24 (1.54–6.82) | 0.002 |
|  |  | 2 | 1.51 (1.27–1.80) | <0.001 | 1.49 (1.23–1.82) | <0.001 | 1.39 (0.83–2.32) | 0.208 | 3.25 (1.55–6.84) | 0.002 |
|  |  | ≥3 | 1.48 (1.07–2.06) | 0.018 | 1.58 (1.11–2.25) | 0.012 | 0.86 (0.26–2.81) | 0.803 | 2.80 (0.78–10.06) | 0.114 |
|  | Male | 1 | 2.46 (2.10–2.88) | <0.001 | 2.40 (2.01–2.86) | <0.001 | 2.48 (1.67–3.70) | <0.001 | 2.83 (1.33–6.04) | 0.007 |
|  |  | 2 | 1.56 (1.31–1.86) | <0.001 | 1.59 (1.30–1.93) | <0.001 | 1.45 (0.92–2.30) | 0.113 | 1.70 (0.70–4.12) | 0.237 |
|  |  | ≥3 | 1.40 (0.97–2.00) | 0.068 | 1.52 (1.04–2.24) | 0.032 | 1.11 (0.40–3.09) | 0.840 | 0.00 (0.00–Inf) | 0.983 |
| Other | All | 1 | 1.46 (1.27–1.67) | <0.001 | 1.33 (1.13–1.57) | <0.001 | 1.88 (1.34–2.63) | <0.001 | 1.93 (1.09–3.42) | 0.025 |
|  |  | 2 | 0.85 (0.74–0.98) | 0.029 | 0.78 (0.67–0.92) | 0.004 | 1.22 (0.87–1.70) | 0.249 | 1.10 (0.61–1.97) | 0.761 |
|  |  | ≥3 | 0.50 (0.38–0.67) | <0.001 | 0.52 (0.38–0.72) | <0.001 | 0.40 (0.17–0.91) | 0.029 | 0.67 (0.23–1.94) | 0.464 |
|  | Female | 1 | 1.34 (1.09–1.66) | 0.006 | 1.24 (0.97–1.59) | 0.083 | 1.87 (1.10–3.18) | 0.021 | 2.21 (0.99–4.93) | 0.054 |
|  |  | 2 | 0.87 (0.71–1.07) | 0.180 | 0.77 (0.60–0.97) | 0.030 | 1.48 (0.91–2.42) | 0.117 | 1.47 (0.67–3.24) | 0.339 |
|  |  | ≥3 | 0.46 (0.30–0.71) | <0.001 | 0.52 (0.33–0.83) | 0.006 | 0.31 (0.07–1.28) | 0.105 | 0.33 (0.04–2.57) | 0.292 |
|  | Male | 1 | 1.57 (1.30–1.90) | <0.001 | 1.44 (1.16–1.79) | 0.001 | 1.91 (1.23–2.96) | 0.004 | 1.70 (0.75–3.89) | 0.206 |
|  |  | 2 | 0.85 (0.70–1.03) | 0.106 | 0.81 (0.65–1.02) | 0.069 | 1.04 (0.66–1.64) | 0.865 | 0.74 (0.30–1.87) | 0.530 |
|  |  | ≥3 | 0.54 (0.37–0.80) | 0.002 | 0.53 (0.34–0.82) | 0.005 | 0.46 (0.17–1.29) | 0.142 | 1.06 (0.30–3.72) | 0.927 |

**Supplementary Table 8.** Marginal derived incidence rates of all strokes per 100,000 person-years by ethnicity, sex, and IMD quintile

|  |  |  | **All strokes** | **Ischaemic Stroke** | **PICH** | **SAH** |
| --- | --- | --- | --- | --- | --- | --- |
| **Ethnicity** | **Sex** | **IMD quintile** | **IRR (95%CI)** | **IRR (95%CI)** | **IRR (95%CI)** | **IRR (95%CI)** |
| White | All | 1 | 107.6 (103.7–112.2) | 83.8 (80.3–87.8) | 12.6 (11.3–14.2) | 5.9 (5.0–7.1) |
|  |  | 2 | 62.7 (59.9–65.6) | 50.4 (48.0–53.3) | 7.4 (6.5–8.5) | 2.7 (2.2–3.5) |
|  |  | ≥3 | 69.3 (63.6–75.0) | 55.2 (50.4–60.1) | 9.8 (7.8–12.0) | 3.0 (2.0–4.6) |
|  | Female | 1 | 100.1 (95.0–106.0) | 76.7 (72.4–82.0) | 11.7 (9.9–13.9) | 6.0 (4.8–8.0) |
|  |  | 2 | 58.6 (54.3–62.8) | 46.0 (42.8–50.0) | 6.6 (5.4–8.2) | 3.1 (2.3–4.5) |
|  |  | ≥3 | 69.0 (62.0–77.9) | 55.2 (48.6–64.1) | 8.5 (6.3–12.0) | 2.9 (1.7–5.5) |
|  | Male | 1 | 114.8 (108.7–121.2) | 90.5 (85.1–96.4) | 13.5 (11.5–16.1) | 5.8 (4.7–7.9) |
|  |  | 2 | 66.4 (62.1–71.1) | 54.5 (50.8–58.6) | 8.1 (6.7–9.9) | 2.3 (1.6–3.4) |
|  |  | ≥3 | 69.9 (62.6–78.1) | 55.6 (48.8–63.7) | 11.0 (8.2–14.9) | 3.1 (1.8–5.5) |
| Black African | All | 1 | 191.4 (176.0–209.0) | 150.5 (136.9–166.8) | 32.0 (26.5–39.1) | 6.2 (4.4–9.1) |
|  |  | 2 | 112.8 (102.7–126.0) | 87.2 (77.7–97.5) | 20.0 (16.0–25.0) | 4.1 (2.7–6.5) |
|  |  | ≥3 | 81.5 (64.5–103.4) | 58.9 (45.5–78.8) | 17.0 (10.4–28.0) | 3.7 (1.4–9.4) |
|  | Female | 1 | 165.5 (144.7–189.1) | 132.4 (115.7–153.7) | 23.9 (17.0–32.8) | 7.1 (4.5–11.8) |
|  |  | 2 | 106.4 (92.7–123.3) | 86.1 (73.3–102.6) | 13.5 (9.1–20.0) | 5.6 (3.4–9.8) |
|  |  | ≥3 | 74.4 (52.0–105.8) | 57.3 (36.5–85.9) | 11.2 (4.7–29.8) | 3.4 (1.0–14.2) |
|  | Male | 1 | 216.6 (193.4–243.9) | 168.1 (148.5–190.2) | 40.1 (32.2–51.5) | 5.1 (2.8–9.6) |
|  |  | 2 | 120.8 (106.3–138.3) | 90.1 (77.6–105.0) | 26.7 (20.2–35.9) | 2.3 (1.0–5.6) |
|  |  | ≥3 | 88.9 (65.7–119.8) | 61.2 (43.7–89.2) | 22.8 (12.4–41.4) | 4.0 (1.0–17.0) |
| Black Caribbean | All | 1 | 152.2 (139.8–165.0) | 118.2 (108.5–129.7) | 22.3 (18.1–27.7) | 9.0 (6.5–12.7) |
|  |  | 2 | 105.7 (96.9–115.5) | 84.3 (76.3–93.6) | 13.7 (10.6–18.0) | 7.5 (5.2–11.3) |
|  |  | ≥3 | 99.9 (78.2–124.6) | 85.6 (68.0–109.4) | 9.6 (4.5–21.5) | 4.5 (1.4–14.8) |
|  | Female | 1 | 135.8 (120.6–153.8) | 106.2 (92.8–121.6) | 18.1 (13.3–25.1) | 9.3 (6.1–14.8) |
|  |  | 2 | 104.4 (92.0–118.3) | 82.3 (72.1–95.5) | 11.9 (8.2–17.8) | 9.4 (5.9–15.7) |
|  |  | ≥3 | 102.4 (76.6–136.6) | 87.1 (62.4–119.6) | 7.3 (2.5–22.0) | 8.1 (2.7–27.4) |
|  | Male | 1 | 172.1 (154.9–190.5) | 133.3 (117.6–150.0) | 27.3 (21.4–36.4) | 8.7 (5.4–15.6) |
|  |  | 2 | 108.9 (96.0–125.1) | 88.1 (76.6–102.7) | 16.0 (11.4–22.9) | 5.2 (2.7–10.7) |
|  |  | ≥3 | 97.7 (69.7–140.5) | 84.8 (58.1–120.2) | 12.2 (5.0–33.0) | 0.0 (0.0–Inf) |
| Other | All | 1 | 100.8 (90.3–112.3) | 73.6 (64.9–83.6) | 18.3 (14.4–24.1) | 5.8 (3.9–8.8) |
|  |  | 2 | 59.2 (53.0–66.2) | 43.2 (37.6–49.3) | 11.9 (9.5–15.3) | 3.3 (2.2–5.2) |
|  |  | ≥3 | 34.9 (26.7–45.7) | 28.9 (21.2–38.8) | 3.9 (1.8–8.4) | 2.0 (0.8–5.9) |
|  | Female | 1 | 92.8 (77.2–111.4) | 68.5 (56.0–84.0) | 16.0 (10.7–23.7) | 6.4 (3.9–11.4) |
|  |  | 2 | 60.0 (51.1–70.7) | 42.2 (34.7–51.3) | 12.7 (9.0–18.3) | 4.2 (2.6–7.3) |
|  |  | ≥3 | 31.9 (21.5–47.9) | 28.9 (18.3–45.6) | 2.6 (0.7–11.5) | 1.0 (0.1–6.8) |
|  | Male | 1 | 110.1 (94.7–128.1) | 79.9 (66.7–96.0) | 21.0 (14.9–29.3) | 5.2 (3.1–10.2) |
|  |  | 2 | 59.5 (51.3–70.8) | 45.2 (38.2–53.8) | 11.5 (8.3–16.3) | 2.3 (1.1–4.8) |
|  |  | ≥3 | 38.1 (27.4–55.6) | 29.2 (18.8–45.2) | 5.1 (2.0–13.1) | 3.3 (1.1–11.8) |

**Supplementary Figure 1.** Changes in ethnic composition and socioeconomic deprivation by ethnicity in the South London Stroke Register catchment population, 1995–2024

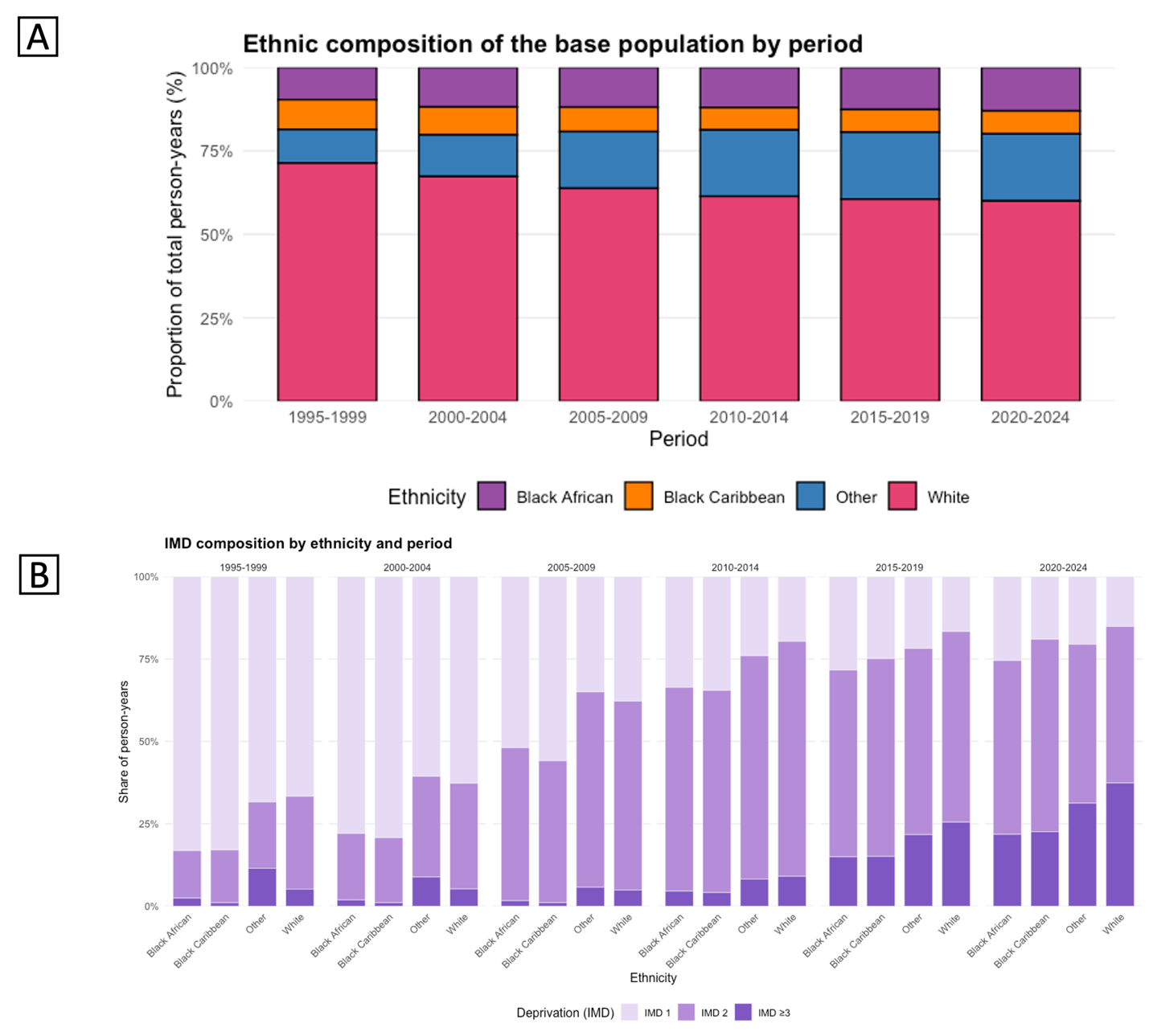

(A) Proportion of total person-years by ethnic group across study periods, showing a decline in the proportion of White residents and growth in Black African, Black Caribbean, and Other groups.

(B) Distribution of IMD categories by ethnic group and period, showing a gradual shift from the most deprived quintile (IMD 1) towards less deprived categories across all ethnic groups.
